# Evaluating the evolution of the timeliness of test-based surveillance systems over the course of a pandemic

**DOI:** 10.64898/2026.02.16.26346417

**Authors:** Ritchie Yu, Philippe N. N. Teichmann, Alina Shimizu-Jozi, Jing Yuan Luo, Rahul K. Arora, Nathan Duarte, Caroline E. Wagner

**Affiliations:** Department of Bioengineering, McGill University, Montreal, Quebec, Canada; Department of Computer Science, McGill University, Montreal, Quebec, Canada; Department of Biology, McGill University, Montreal, Quebec, Canada; Department of Information Technology and Electrical Engineering, ETH Zurich, Zurich, Switzerland; Cumming School of Medicine, University of Calgary, Calgary, Alberta, Canada; Department of Electrical and Computer Engineering, McGill University, Montreal, Quebec, Canada

## Abstract

The timeliness of infectious disease surveillance systems largely determines the speed at which mitigation interventions may be implemented. However, it is unclear how surveillance timeliness evolves during a pandemic with changing government policies, testing tools, and population-level infection and immunity landscapes. Here, we adapt an agent-based model for COVID-19 transmission to explore the timeliness of the surveillance signals obtained from polymerase chain reaction (PCR) and rapid antigen (RAT) tests relative to true infection incidence. Across different pandemic scenarios, we investigate how surveillance timeliness depends on the prevalence of co-circulating influenza-like-illnesses (ILI) and test quality. If only PCR tests are available with symptom-based eligibility, and if tests can detect post-recovery residual viral load, then a surveillance lag may emerge which is amplified by ILI prevalence. When limited RATs are introduced with symptom-based eligibility, and PCR eligibility requires a recent positive RAT, then RAT/PCR timeliness is sensitive to ILI prevalence but insensitive to RAT failure probability. With unrestricted RAT supply, PCR timeliness varies with both ILI prevalence and RAT failure probability. Our work highlights how the timeliness of test-based surveillance signals can evolve throughout a pandemic, with important implications for interpreting real-time surveillance data and designing more effective, data-driven surveillance systems.

## 2 Introduction

Effective surveillance systems are needed to guide robust public health responses during a pandemic arising from the emergence of a novel pathogen. The aim of surveillance is to estimate a population’s health status by collecting and analyzing health-related data, in order to guide the deployment of interventions and measure their impact [1]. Here, we focus on test-based surveillance systems, which aggregate results from diagnostic tests and were commonly used during the COVID-19 pandemic. Note that despite their use in surveillance, the primary purpose of a diagnostic test is not surveillance, but rather to identify whether a patient has a disease to guide treatment [2]. Due to this reliance on routinely collected testing data, test-based surveillance systems are passive systems [3].

Timeliness is a critical feature of an effective surveillance system [4]. Timeliness can refer to the delay between different steps of a surveillance system, such as the time from the onset of an infection to the infection being reported to a public health agency [4]. Timeliness can also refer to the delay between a surveillance signal and some pandemic signal. Especially for pandemics arising from rapidly-spreading infectious diseases, a timely surveillance system can be valuable to prompt intervention measures earlier for mitigating disease spread [1, 5]. By calibrating a disease spread network, Najeebullah et al. found that for dengue outbreaks in Australia, shorter surveillance response delays would lead to larger reductions in epidemiological indicators such as outbreak size and number of cases prevented [6]. Substantially smaller reductions were expected to be obtained as the response delay increased, which emphasizes the importance of timely surveillance for more effective disease mitigation [6]. Note however that the level of timeliness needed for disease mitigation can vary across different infectious disease due to differences in the speed of disease progression [7]. The importance of timeliness is also evident in test-based population screening aimed at identifying infectious individuals to break transmission chains. Larremore et al. showed that for the COVID-19 pandemic, it was more important to improve reporting delays and test frequency, rather than test sensitivity, as improving the latter led to more marginal improvements in screening effectiveness [8]. While timeliness is a crucial feature of effective surveillance systems, optimizing for timeliness can sometimes come at the expense of other surveillance attributes. For test-based surveillance systems, Cilloni et al. highlighted the trade-off between rapid isolation afforded by lateral flow assays and the reduction of unnecessary isolations due to false positives offered by confirmatory polymerase chain reaction (PCR) testing [9].

Pandemics are highly dynamic and often exhibit significant evolution in multiple important variables, which include the state of available testing technologies as well as the epidemiological and immunological states of the populations in question. For instance, during the COVID-19 pandemic in Quebec, Canada, diagnostic test results provided a commonly used surveillance signal to estimate the true incidence of infection [10]. Early in the pandemic, when overall infection levels were relatively low, tests in Quebec mainly comprised of PCR tests. In May 2020, during the first major wave of infections, Quebec had around 6000 daily PCR tests reserved for symptomatic individuals [11, 12]. PCR tests are the gold standard for COVID-19 diagnosis, and focus on amplifying viral RNA in mucus samples, which are often collected through nasal swabs and then transported to a laboratory for analysis [13]. Globally, eligibility criteria for receiving a PCR test was generally stringent at the start of the COVID-19 pandemic, with priority allocated towards individuals with COVID-19-like symptoms. Although eligibility criteria relaxed as test capacity improved over time, resource limitations posed a significant obstacle for the push to scale PCR testing. Over time, alternative testing tools which were faster and cheaper to administer became increasingly developed and relied upon, most notably rapid antigen tests (RATs) [14]. RATs are lateral flow assays designed to detect SARS-CoV-2 antigens present in patient mucus samples, often collected via nasal swab. The widespread adoption of RATs in Quebec was delayed due to caution and prudence, as health officials were concerned with limitations in test accuracy [15–17]. For example, while the federal government delivered 2.6 million rapid screening tests to Quebec in October 2020, including 1.2 million RATs, only 24,000 had been used by early February 2021 [16, 17]. However, by late December 2021, coinciding with the emergence of the Omicron variant, RATs became widely and freely distributed through pharmacies in Quebec, as 4.3 million RATs were planned to be released in the first week of the new pharmacy distribution strategy [18–20]. While test quantities distributed through pharmacies were free, their supply was still restricted as individuals over age 14 could only obtain a 5-test kit every 30 days [18].

Other factors also challenge the ability to maintain surveillance timeliness throughout an outbreak of a specific pathogen. For example, PCR tests and RATs fundamentally differ in their turnaround time: it typically takes 1-3 days for a PCR test result to be returned, while RAT results can be available within 15-30 minutes [21]. Therefore, surveillance timeliness may vary depending on the relative quantity of each type of test used to conduct surveillance. During an outbreak, there may also be co-circulating pathogens that illicit similar symptoms. For COVID-19, infection with a respiratory pathogen other than SARS-CoV-2 (which we broadly refer to here as “influenza-like-illness” or ILI) may prompt individuals without COVID-19 to seek a test. When ILI incidence is sufficiently high, one may hypothesize a rise in testing pressure which may worsen surveillance timeliness, for example by increasing test result turnaround time due to slowed laboratory processing or insufficient staffing. Indeed, periods of heightened testing demand (not necessarily linked to increased ILI incidence) worsened test turnaround time during the COVID-19 pandemic in Quebec [22]. Any impact of ILI incidence on test consumption patterns would likely vary as ILI incidence follows historically established seasonal trends, though these trends may not necessarily hold during an outbreak. [23, 24]. During the COVID-19 pandemic, seasonal trends for ILI incidence deviated from historical trends as a consequence of non-pharmaceutical interventions (NPIs) which aimed to reduce COVID-19 spread [25]. Furthermore, co-circulating pathogens have the potential to interact with each other, meaning that one pathogen may influence the risk of infection with another [26]. These types of interactions at the host level can affect the incidence of each pathogen at the population level, due to changes in parameters such as infection susceptibility and transmissibility [26–28].

The timeliness of a collected surveillance signal is ideally measured against the true incidence of infection, yet the latter variable is generally impossible to accurately measure since it requires identifying infections almost as soon as they arise. Case identification during the COVID-19 pandemic was also challenged by imperfect test accuracy, limited test capacity, and the presence of individuals who were asymptomatic or had mild symptoms [29–31]. Therefore, a common approach is to retrospectively compare a surveillance signal against a reference signal which is simpler to measure, for example hospital admission rates [32, 33]. However, these analyses assume that the reference signal is an accurate substitute for the true incidence of infection, which is not always valid. [34, 35]. For example, the signal obtained from hospital admission rates may be biased by temporal changes in health-seeking behaviour [34]. Infection prevalence obtained through random sampling [36] can serve as a less biased reference signal, but may still be imperfect due to individuals with long-term SARS-CoV-2 RNA shedding [34, 37]. Consequently, mathematical models are powerful tools for evaluating surveillance system effectiveness as a result of their ability to simulate an accessible baseline incidence of infection along with how this signal responds to various interventions. For example, Smith et al. attempted to optimize COVID-19 surveillance within long-term care facilities [38]. Using an agent-based model (ABM), they explored different PCR testing strategies and observed that the time to detect an outbreak depended on test capacity and the specific testing strategy [38]. However, most other modeling studies focus on how successful interventions are for averting infections [39–41]. As a result, additional theoretical work is needed to adequately characterize surveillance system effectiveness, and how it is impacted by evolving technological and epidemiological conditions during a pandemic.

Here, we adapt a previously developed ABM for COVID-19 [39] to study the timeliness of both PCR-based and RAT-based testing surveillance systems under four different scenarios that roughly correspond to historical “eras” of the COVID-19 pandemic. These eras are characterized by the quality and quantity of testing technologies, the criteria in place for being test-eligible, as well as the epidemiological landscape. Overall, our work contributes to establishing conditions under which specific types of test-based surveillance systems may be more or less reliable, supporting the establishment of targeted, responsive, data-driven, and effective public health responses in settings of disease transmission.

## 3 Methods

### 3.1 Baseline disease modeling framework

We use an ABM framework as these models lend themselves to simulating test-based surveillance systems. Particularly, test eligibility criteria can be specified to arbitrary detail, and eligibility can be evaluated by checking individual agent attributes. In contrast, the graph associated with a compartmental model can quickly become complex and difficult to manage as the number of compartments increases. We adapt the previously-published open-source ABM Covasim [39] to simulate COVID-19 transmission. Covasim simulates populations of individual agents linked through contact networks, and agents are assigned disease states based on these interactions (i.e., susceptible, exposed, infectious, recovered, or dead). The Covasim population network can be generated as a random network, hybrid network, or a SynthPops network. SynthPops is a package used to build realistic synthetic population networks [39]. Different contact layers model agent interactions within different settings that include school, household, workplace, and community [39]. For a given simulation, initial infections are seeded and then, over a number of discrete time steps, Covasim uses a pseudorandom number generator to determine agent transitions between states. The probability of disease transmission from source agent *p*_1_ to target agent *p*_2_ depends on the population setting in which contact takes place, as well as the relative transmissibility of *p*_1_ and the relative susceptibility of *p*_2_. Both relative transmissibility and relative susceptibility are age-dependent, but also depend on the agent’s immediate context, such as their viral load, immunity, and whether they are quarantined or isolated.

We use a custom variation of SynthPops (see Supplementary Material) to generate a population containing *N* _agent_ = 20, 000 agents, parameterized by demographic distributions (e.g., age, school enrollment rate, household size) from the United Kingdom. When data could not be found for certain population fields, we relied on existing United States data as an approximation (see Supplementary Material). Each simulation is initialized with 30 seed infections and each simulation lasts 400 time steps (days). When an agent receives a positive test (PCR test or RAT), they enter the “diagnosed” state, and we assume that all of the agent’s contacts are immediately notified of their close contact with a confirmed case. Close contacts are then eligible to receive a test if the associated eligibility criterion is enabled, but they do not make behavioural changes such as quarantine or isolation. We also make the important assumption that an agent taking a test does not make any behavioural change in response to the test result. This assumption eliminates the ability of tests to impact transmission, allowing us to focus our analysis solely on how the temporal surveillance signal associated with different testing conditions compared to the underlying infection curve.

### 3.2 Modifications to Covasim

#### 3.2.1 Background influenza-like-illness

We incorporate a simple model of background ILI into the Covasim framework, wherein a proportion *P*_ILI_ of the original population size (discounting any deaths) is constantly infected with ILI throughout the simulation. Each individual recovers within *t* ∼ *𝒰* (7, 10) days, in other words *t* is an integer sampled uniformly between 7 and 10 days inclusive. We assume all agents with ILI are symptomatic throughout the duration of their illness. Note that in contrast, agents with COVID-19 are often not symptomatic for the full duration of their illness, since they either have symptoms arising with some randomly sampled delay, or they have an asymptomatic infection. Each day, ILI recoveries are checked and new infections are randomly distributed among the population that is not already infected with COVID-19 or ILI to maintain *P*_ILI_. With the incorporation of ILI, we aim to simulate a scenario where an infection has been circulating for a prolonged period of time prior to the onset of a pandemic with a different pathogen. To model this, at the start of the simulation, we transform an initial recovery day distribution with dates *t* ∼ *𝒰* (7, 10) and assign the resulting recovery day distribution to agents selected to have ILI on day 0 of the simulation. By applying this procedure, we ensure that recovery dates are well-mixed during the early days of the simulation, which is more representative of a pathogen that has been previously co-circulating (see the Supplementary Material and Figure S1 for details).

#### 3.2.2 Viral load

By default, Covasim models viral load kinetics as a step function that switches from high to low viral load at some transition point during an infection. We implement a more realistic viral load model to incorporate *LOD*-based (compared to probabilistic) testing. We generally follow the principles outlined by Larremore et al. to model viral load with (i) a variable latent period, (ii) a rapid growth phase from low to peak viral load, (iii) a slower decay phase, and (iv) longer clearance for symptomatic infections as opposed to asymptomatic infections [8]. Specifically, we model viral load trajectories as continuous linear piecewise functions [8]. When an agent becomes infected with COVID-19, they are assigned anchor points corresponding to different disease stages, and linear interpolation between anchor points produces the agent’s full viral load trajectory. The parameters used to define these anchor points (see below) were chosen by inspection for their ability to generate viral load kinetics featuring properties (ii) and (iii) [8] (see Figure S2). Meanwhile, Covasim natively models (i) by stochastically sampling latent periods (time from exposure to infectiousness) for each infection, and accommodates (iv) by modeling prolonged recovery of symptomatic agents relative to asymptomatic agents.

When an agent is infected with COVID-19, we compute the following anchor points. We first compute the point (*x*_1_, *y*_1_) where viral load reaches *y*_1_ = 3 at time *x*_1_. Note that in our algorithm, given a point (*x, y*), the value *y* is always logarithmic and refers to an actual viral load of 10^*y*^ cp/ml. We compute *x*_1_ as

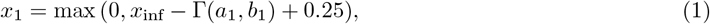

where *a*_1_ = 2 and *b*_1_ = 0.35 are shape and scale parameters of the Gamma distribution. Additionally, *x*_inf_ is the time at which an agent becomes infectious in Covasim, which is known before computing *x*_1_. Using *x*_inf_, the second point in our model is then (*x*_inf_, *y*_inf_), where logarithmic viral load reaches *y*_inf_ = 6 at time *x*_inf_. The third point in our model is (*x*_2_, *y*_2_), where logarithmic viral load achieves a peak value of *y*_2_ at time *x*_2_. We compute *x*_2_ from

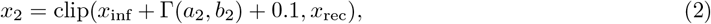

where *a*_2_ = 3 and *b*_2_ = 0.26 are again shape and scale parameters of the Gamma distribution. The term *x*_rec_ is the agent’s recovery date, which is known before computing *x*_2_, and we clip *x*_2_ such that it cannot exceed *x*_rec_. Note that a small number of agents may die, and in these cases we replace *x*_rec_ with the date of death. We compute the peak *y*_2_ as

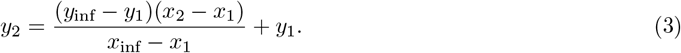

The final point is (*x*_rec_, *y*_rec_), where logarithmic viral load reaches *y*_rec_ = 6 at predetermined time *x*_rec_. Between time *x*_inf_ (inclusive) and *x*_rec_ (exclusive), we say an agent is infectious to follow the naming convention used by Covasim [39]. Using the defined anchor points, on any given day *t* we perform linear interpolation to calculate an agent’s logarithmic viral load *y*_*t*_ as

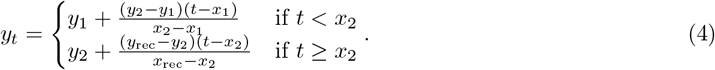

Viral loads which fall below 10^0^ cp/ml are clipped to 0 cp/ml. Note that despite the definition of (*x*_rec_, *y*_rec_) above, in practice when time *x*_rec_ is reached, we replace *y*_rec_ with *y*_rec_ − *ϵ* where *ϵ* = 10^*−*5^. This ensures that an agent is in the infectious state if and only if they have viral load *y* ≥ 6. Sample viral loads produced by this model for 1000 agents are illustrated in Figure S2.

#### 3.2.3 Diagnostic testing framework

We built a custom testing pipeline which on a daily basis decides which agents are test-eligible, then test-seeking, and finally granted a test followed by receipt of the test result with potential delay. Built on our models of viral load and background ILI, our testing framework also incorporates *LOD*-based testing and, with appropriately specified eligibility criteria, permits the usage of tests for reasons other than having COVID-19. While Covasim’s native implementation provides a default testing framework, a custom framework was required to more comprehensively model the above features. First, in Covasim’s implementation, agents are assigned a probability of taking a test which is updated if they undergo certain state transitions (e.g., become symptomatic or enter quarantine). Our approach allows us to precisely index which agents are eligible and eventually take a test. Second, test results in Covasim are not *LOD*-based, and are instead determined through parameters for test sensitivity and specificity [39]. Our approach introduces *LOD*-based testing by considering an agent’s time-varying viral load. Third, while Covasim considers the effect of co-circulating ILI directly within their testing framework, their approach re-samples ILI infections each day, and hence does not model continuity of ILI infection within individual agents. Our approach builds upon our model of ILI circulation, which considers continuity of infection and thus enables more realistic behaviours such as test-seeking across several consecutive days. Finally, we note that while our approach and Covasim’s approach both allow for different tests (e.g., PCR and RAT) to be simultaneously deployed in the population, our ability to precisely model test eligibility opens the door for interactions between testing systems (e.g., PCR test-seeking prompted by a positive RAT).

##### Test attributes and definitions

Prior to the start of a simulation, each type of test (PCR or RAT) is associated with a set of eligibility criteria. Each criterion is associated with a fixed probability of test-seeking that is constant across agents. In the absence of behavioral data to parameterize test-seeking probabilities, we set all probabilities to *P*_test_ = 0.5. Each type of test is also assigned attributes listed in Table 1.

**Table 1.** Description of attributes assigned to PCR tests and RATs.

| Attribute | Symbol | Description |
| --- | --- | --- |
| Daily test capacity | $N$ | Daily maximum number of tests that may be allocated and used in the population |
| Limit of detection | $LOD$ | Lowest detectable concentration of viral load (cp/ml) expressed on a logarithmic scale |
| Failure probability | $P_{\text{fail}}$ | Probability that a test fails and returns the opposite result |
| Positive re-test delay | $T_+$ | Number of days an agent must wait to test again after taking a will-be-positive test |
| Negative re-test delay | $T_-$ | Number of days an agent must wait to test again after taking a will-be-negative test |
| Reporting delay | $T_r$ | Number of days before a test result becomes known to an agent |

A test result returns positive if the agent’s viral load is greater than or equal to the limit of detection, and returns negative otherwise. With probability *P*_fail_, the test fails and returns the opposite result. Test results are returned to agents with a delay of *T*_*r*_ days. If an agent’s test returns positive, then they cannot take a test of the same type for *T*_+_ days after the original test date. Similarly, a negative test prohibits an agent from taking a test of the same type for *T*_*−*_ days after the original test date. Note that in our implementation, it is not possible for an agent’s test result to be lost.

##### Test allocation algorithm

On a given day, we first identify which agents in the population are eligible for testing according to the eligibility criteria. Whether an agent seeks a test on the basis of one criterion is independent of other criteria. Therefore, if an agent satisfies *n* criteria, the probability they do not seek a test is (1 − *P*_test_)^*n*^. Once we have identified test-seeking agents, we then allocate tests based on the current day’s test capacity *N*. If *N* exceeds the number of test-seeking agents, then all test-seeking agents will be granted a test. Unused tests are not carried over to the next day. Otherwise, if test capacity is insufficient, then given test eligibility criteria {*C*_1_, *C*_2_, …, *C*_*M*_}, we identify all possible subsets of criteria, excluding the empty set. For each subset of criteria, we identify the corresponding group of agents who precisely satisfy such criteria. Tests are then distributed proportionally to the size of subset groups. If *s* agents belong to a subset group, and there are *t* eligible agents in total, then 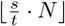 tests are reserved for the subset group. Due to rounding, a few tests may go unused. In this case, each remaining test is randomly distributed to a subset group, where the probabilities are proportional to subset group size.

##### Agent response to test result

A key simplification we make is to assume that test results do not cause agents and their close contacts to make any behavioral adjustments. The one exception is potential test-seeking among close contacts if they subsequently satisfy eligibility requirements. We do this to isolate the effect of our specific parameters of interest on the timing of the positive test curve relative to the baseline infection signal, without the latter being modified by quarantine/isolation adherence. To implement this assumption, we set Covasim’s isolation factors to 1.0, such that a positive result does not reduce agent transmissibility. Furthermore, we disable all quarantine behaviour in Covasim and set all quarantine factors to 1.0, to prevent any quarantine-related impact on agent transmissibility/susceptibility.

It is important to note that in the context of this work, the low assumed LOD of the PCR tests results in the possibility of a detectable viral load (producing a positive PCR test) after an agent has recovered, since agent recovery coincides with when their logarithmic viral load value falls below 6. This phenomenon was shown to occur in the real-world COVID-19 pandemic, as recovered patients were observed to receive positive PCR tests [42]. For consistency, we define “true positive” and “true negative” test results as those arising when the test returns the correct status based on the detected viral load. For instance, a positive test for an agent who has recovered but has a logarithmic viral load greater than the *LOD* is considered a true positive. In this way, false positives and false negatives only arise due to test failure, which occurs probabilistically at rate *P*_fail_ and may be different between PCR tests and RATs.

#### 3.3 Measuring surveillance timeliness

For a simulation lasting *d* days, suppose ***x*** = (*x*_0_, …, *x*_*d*−1_) is the daily number of new infections (i.e., incidence) and ***y*** = (*y*_0_, …, *y*_*d*−1_) is the daily number of new positive test results. We standardize the signals by setting

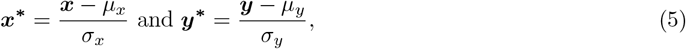

where *µ*_*x*_, *µ*_*y*_ are the means of ***x, y*** respectively, and *σ*_*x*_, *σ*_*y*_ are the standard deviations of ***x, y*** respectively. We then compute *τ*, the number of days by which ***y*** lags ***x*** as

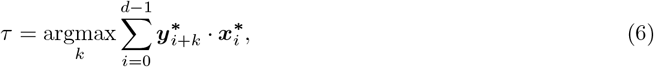

where *k* ∈ {−*d* + 1, −*d* + 2, …, *d* −1}. We compute each summation with zero padding, meaning that if *i* + *k* < 0 or *i* + *k* > *d* − 1, then we set 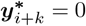. We compute cross-correlation with zero padding as well as the argmax in Equation 6 using the NumPy library in Python.

### 3.4 Pandemic scenarios

We explore several pandemic scenarios loosely corresponding to different chronological “eras” during the COVID-19 pandemic. Each scenario is characterized by a set of fixed variables defined in Table 2. Within each scenario, we sweep certain variables of interest and identify trends in how test-based surveillance systems perform in response. Note that all experiments are performed with fixed population structure and transmission characteristics. If these are modified, then the variables in Table 2, such as the number of available daily tests, would likely need to be tuned to observe interpretable trends.

**Table 2.** Eligibility criteria and test attributes used for Scenarios A, B, C, and C^*^. An agent can be “symptomatic” with COVID-19 or ILI (Scenarios A, B, C, C^*^). An agent with COVID-19 may progress into a “severe/critical” disease state, where progression probability is age-linked (Scenario A) [39]. If an agent contacted a confirmed case on the previous day, then they satisfy “contact with confirmed case” (Scenario B). If an agent contacted a symptomatic agent on the current day, then they satisfy “contact with symptomatic agent” (Scenario B). The “other” eligibility criterion is meant to model other eligibility criteria we have not accounted for (Scenario B). Each day, 1% of the population is randomly selected to satisfy the “other” criterion. For the “workplace screening” criterion, at the start of each simulation we randomly select 5% of workplaces to participate in workplace screening, where each employee performs a test every 7 days (Scenario B). Exceptionally, agents who participate in workplace screening are not restricted by *T*_+_ and *T*_*−*_ re-test delays. An agent is eligible to receive a PCR test if they received a positive RAT in the previous 3 days (Scenarios C, C^*^). In Scenarios A and B, we assume *N* = 300, *T*_*r*_ ∼Γ(3, 1.5), and *T*_*−*_ = 14 for PCR tests. We set the PCR failure probability to *P*_fail_ = 0.01 as an approximation of the known high sensitivity and specificity of PCR tests [43, 44]. We set *T*_+_ = 90 based on CDC guidance that PCR tests may continue to return positive 90 days after an initial positive test [45]. However, note that our re-test delay period is defined following the original test date, whereas CDC guidelines begin the re-test delay period following receipt of the test result. In Scenario C, we assume daily test capacity for RATs to be *N* = 600, while in Scenario C^*^, we assume *N* = 20000 such that it is possible for the entire population to take a RAT on any given day. In both Scenarios C and C^*^, we set *LOD* = 6 for RATs so that a recovered agent will never receive a true positive. This choice is further justified in the Scenario C subsection of the Results section. We assume *T*_+_ = 14, *T*_*−*_ = 0, and *T*_*r*_ = 0 for RATs. In both Scenarios C and C^*^, the assumptions for PCR test attributes are similar to Scenarios A and B. However, since we are now sweeping *P*_fail_ for RATs, we fix *LOD* for PCR tests at *LOD* = 6, again to ensure that a recovered agent will never receive a true positive PCR test when they are not infectious.

| Scenario | Test | Eligibility criteria | $N$ | $LOD$ | $P_{\text{fail}}$ | $T_+$ | $T_-$ | $T_r$ |
| --- | --- | --- | --- | --- | --- | --- | --- | --- |
| A | PCR | + Symptomatic<br>+ Severe/critical | 300 | varied | 0.01 | 90 | 14 | $\Gamma(3, 1.5)$ |
| B | PCR | + Symptomatic<br>+ Contact with confirmed case<br>+ Contact with symptomatic agent<br>+ Other<br>+ Workplace screening | 300 | varied | 0.01 | 90 | 14 | $\Gamma(3, 1.5)$ |
| Scenario | Test | Eligibility criteria | $N$ | $LOD$ | $P_{\text{fail}}$ | $T_+$ | $T_-$ | $T_r$ |
| C | RAT | + Symptomatic | 600 | 6 | varied | 14 | 0 | 0 |
| | PCR | + Received positive RAT in last 3 days | 300 | 6 | 0.01 | 90 | 14 | $\Gamma(3, 1.5)$ |
| C* | RAT | + Symptomatic | 20000 | 6 | varied | 14 | 0 | 0 |
| | PCR | + Received positive RAT in last 3 days | 300 | 6 | 0.01 | 90 | 14 | $\Gamma(3, 1.5)$ |

First, we explore two scenarios (A and B) where testing is limited to PCR testing, approximately corresponding to earlier stages of the pandemic when RATs were not broadly available. Scenario A uses a more stringent set of criteria for test eligibility compared to Scenario B, corresponding to the relaxation of eligibility criteria over time. In Scenarios A and B, we explore how surveillance system performance depends on *P*_ILI_ and the *LOD* of PCR tests. We sweep *P*_ILI_ in the interval [0.004, 0.800], and sweep the *LOD* for PCR tests in the interval [2.0, 6.0]. We then explore two scenarios (C and C^*^) where testing includes both PCR tests and RATs. RATs emerged later in the pandemic, and were an important diagnostic tool when high costs and a lack of trained personnel and test reagents constrained the ability of countries to sufficiently scale up PCR testing [14]. In Scenario C^*^, we assume that RAT capacity is sufficiently high such that the entire population could obtain a test on a given day, whereas in Scenario C, RAT availability is limited to 600 tests daily. In both scenarios, we maintain PCR test capacity at 300 tests daily. In Scenarios C and C^*^, we explore how surveillance system performance depends on *P*_ILI_ and *P*_fail_ for RATs. We sweep *P*_ILI_ in the interval [0.004, 0.800], and sweep *P*_fail_ for RATs in the interval [0.00, 0.50]. Within each scenario, for a given pair of variable values, we run the corresponding simulation 100 times with a different random seed each time. For each simulation, surveillance timeliness is calculated using the method previously described. We then aggregate the timeliness values into a single mean representation. Note that for attributes which are a function of time in days, such as daily test consumption, we take the mean attribute value for each day across 100 independent simulations.

## 4 Results

### 4.1 Scenario A

We begin by simulating surveillance timeliness in Scenario A, where only PCR tests are available and test eligibility is stringent, with symptoms or a severe/critical disease state being required to seek a test. This scenario is designed to correspond to the early stages of a pandemic, when diagnostic tools are limited. In Figure 1**A**, we plot surveillance timeliness as a function of both the PCR test *LOD* (where a **lower** *LOD* corresponds to a **more** sensitive test) and the background level of ILI.

**Fig 1.**
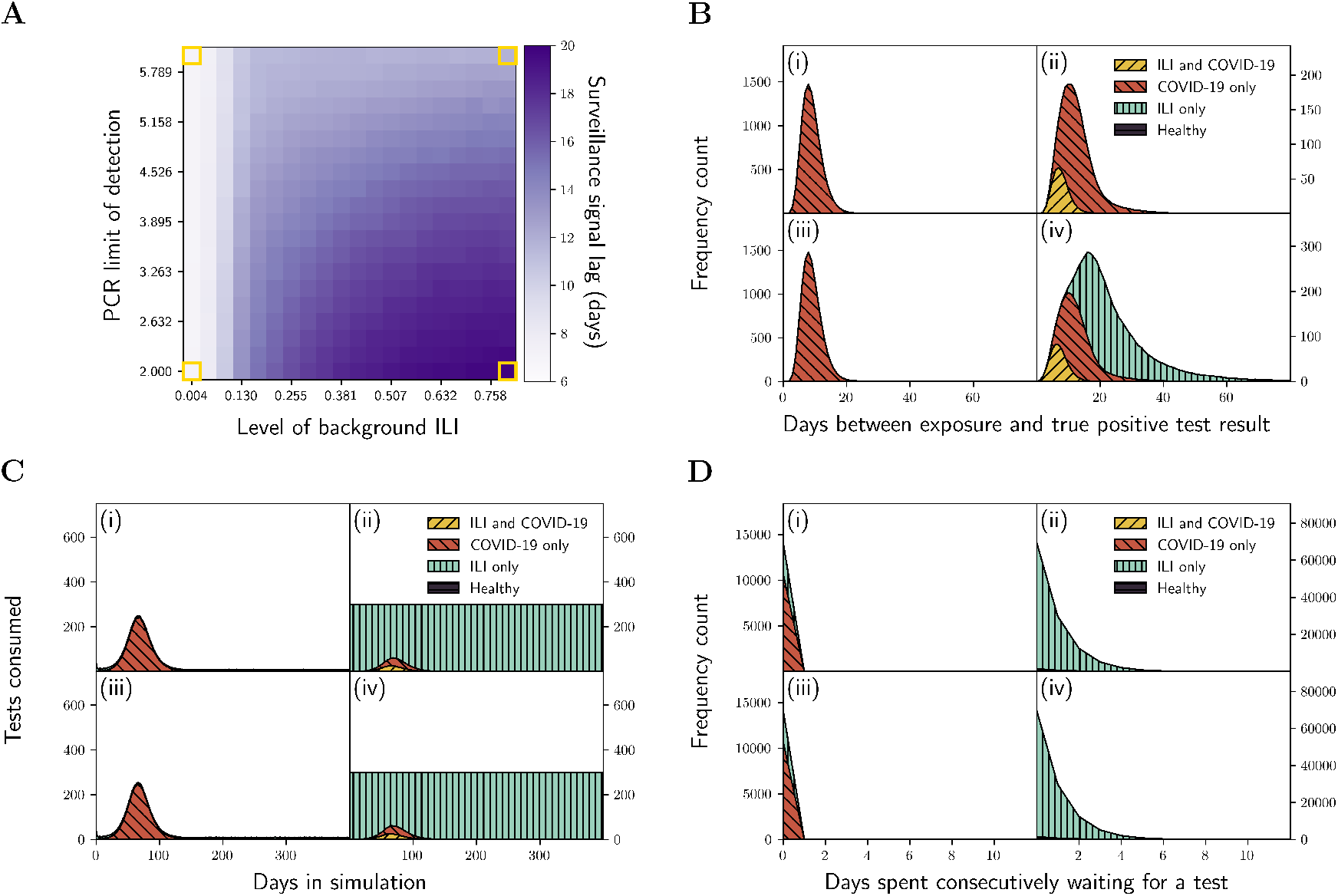
**A:** Surveillance timeliness in Scenario A as a function of PCR test *LOD* and *P*_ILI_. A positive timeliness value indicates that the surveillance signal (daily positive PCR tests) lags the pandemic signal (daily new infections). Yellow highlighted boxes at the heatmap corners identify settings of (*P*_ILI_, *LOD*) used to construct the corresponding quadrants in the **B, C**, and **D** subplots. **B:** Distribution of time between when an agent becomes exposed to COVID-19 to when they receive a true positive test result. The quadrant settings are (i) (*P*_ILI_, *LOD*) = (0.004, 6), (ii) (*P*_ILI_, *LOD*) = (0.800, 6), (iii) (*P*_ILI_, *LOD*) = (0.004, 2), and (iv) (*P*_ILI_, *LOD*) = (0.800, 2). Furthermore, for each quadrant, the curve is partitioned by infection status at the time of testing, each associated with a unique color/hash combination. **C:** Daily number of PCR tests consumed. **D:** Distribution of the number of days spent consecutively seeking a PCR test before receiving one. The quadrant settings and color/hash schemes in **C** and **D** are identical to those in **B**.

We find that in general, timeliness worsens at low PCR test *LOD* and high *P*_ILI_. This somewhat counter-intuitively implies that a more sensitive PCR test can reduce surveillance effectiveness under certain conditions. To understand the origin of this finding, in Figure 1**B**, we plot the distribution of times between when an agent is exposed to COVID-19 and when they receive a true positive PCR test result for different agent infection categories. The category that an agent belongs to is determined at the time of the PCR test. We plot these distributions for four pairs of background ILI level / PCR test *LOD* corresponding to the highlighted squares in the four corners of Figure 1**A**.

A long tail emerges in Figure 1**B**(iv) associated with agents currently symptomatic with an ILI infection (the green shaded region), but who retain a detectable residual viral load from a previous COVID-19 infection despite no longer being infectious with COVID-19. The long tail disappears in Figure 1**B**(ii) at identical *P*_ILI_ but higher PCR test *LOD*, since the PCR test is no longer sensitive enough to detect residual viral load. In our simulations, we also monitor the number of consecutive days an agent spends test-seeking before receiving a test. During periods of high test demand, agents may need to wait several days before receiving a test as finite test capacity forms a bottleneck. We term the shortage of tests “supply crowding” and the need to wait for a test “temporal crowding”. These terms become critical later to explain the results we obtain in Scenarios C and C^*^, but are introduced here for consistency. In Scenario A, we observe supply crowding at high *P*_ILI_, as test capacity *N* = 300 is met essentially every day (Figure 1**C**(ii) and Figure 1**C**(iv)). Correspondingly, we observe temporal crowding at high *P*_ILI_ (Figure 1**D**(ii) and Figure 1**D**(iv)), as many individuals with only ILI wait several days to receive a PCR test. The waiting time distribution corresponding to the “COVID-19 only” infection status also broadens in Figures 1**D**(ii) and 1**D**(iv) compared to Figures 1**D**(i) and 1**D**(iii) (see Figure S4). We highlight that while the waiting time distribution remains similar between Figure 1**D**(ii) and Figure 1**D**(iv), timeliness is comparatively worse in Figure 1**D**(iv) due to the detection of residual viral load which is only possible at low test *LOD* (see Figure 1**A**).

### 4.2 Scenario B

We repeat this exercise for Scenario B, which has more relaxed test eligibility criteria compared to Scenario A. This loosely represents a later pandemic “era” in which PCR tests have become more accessible, yet PCR test capacity is still finite and RATs still remain unavailable. As seen in Figure 2**A**, surveillance timeliness once again generally worsens for lower PCR *LOD*, but is relatively insensitive to background ILI levels. As seen in Figure 2**B**, the timeliness trend with PCR *LOD* arises once again as a result of the detection of residual viral load long after recovery from a previous COVID-19 infection (compare the long tails in Figures 2**B** (iii) and (iv) with the tighter distributions in Figures 2**B** (i) and (ii)). Unlike in Scenario A, however, these delayed true positive tests for low PCR *LOD* can also arise in currently healthy individuals, since several test eligibility criteria in Scenario B such as “contact with confirmed case” may be satisfied by an individual without an ongoing ILI or COVID-19 infection. Consequently, PCR test timeliness is impacted for all background levels of ILI in this scenario.

**Fig 2.**
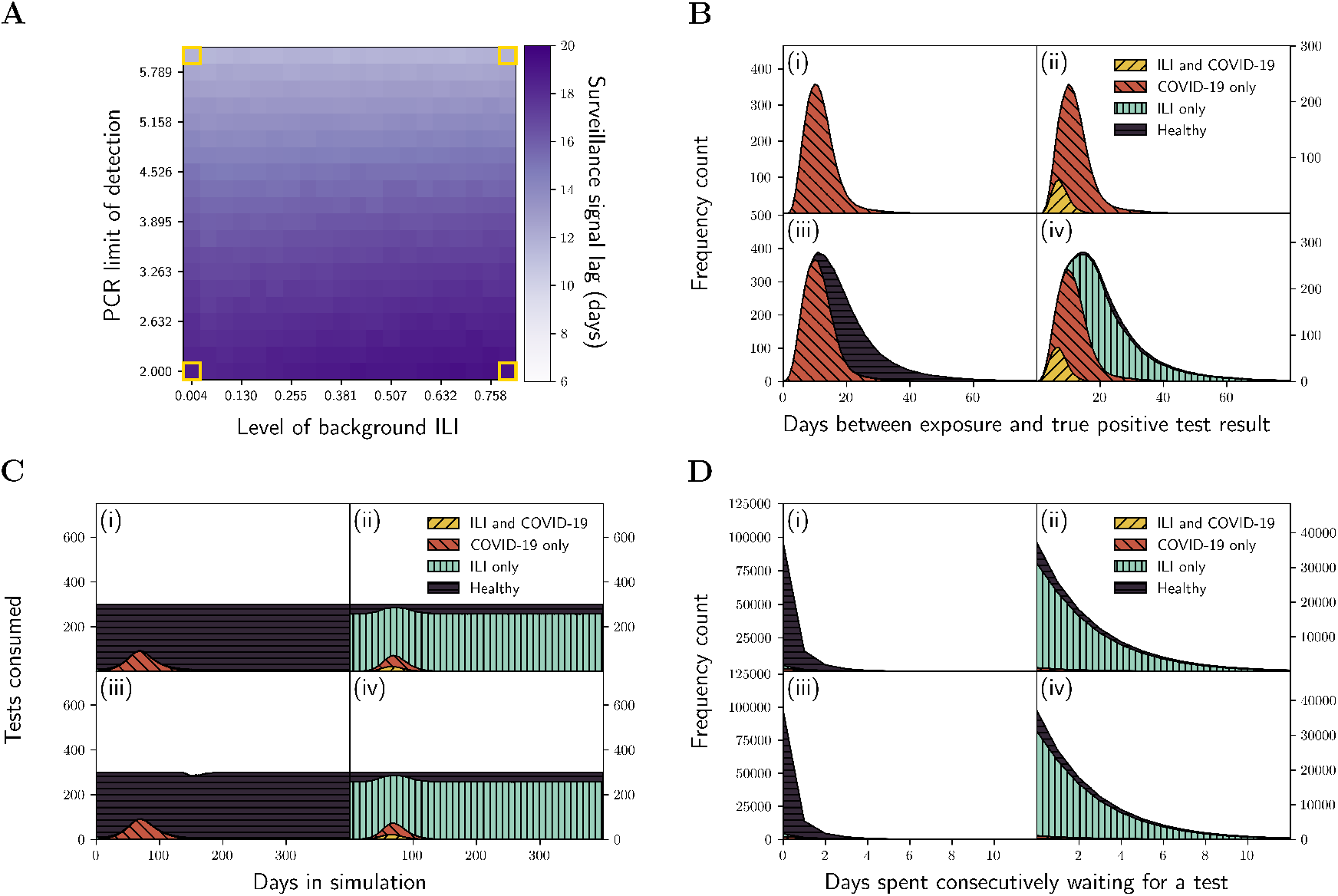
**A:** Surveillance timeliness in Scenario B as a function of PCR test *LOD* and *P*_ILI_. A positive timeliness value indicates that the surveillance signal (daily positive PCR tests) lags the pandemic signal (daily new infections). Yellow highlighted boxes at the heatmap corners identify settings of (*P*_ILI_, *LOD*) used to construct the corresponding quadrants in the **B, C**, and **D** subplots. **B:** Distribution of time between when an agent becomes exposed to COVID-19 to when they receive a true positive test result. The quadrant settings are (i) (*P*_ILI_, *LOD*) = (0.004, 6), (ii) (*P*_ILI_, *LOD*) = (0.800, 6), (iii) (*P*_ILI_, *LOD*) = (0.004, 2), and (iv) (*P*_ILI_, *LOD*) = (0.800, 2). Furthermore, for each quadrant, the curve is partitioned by infection status at the time of testing, each associated with a unique color/hash combination. **C:** Daily number of PCR tests consumed. **D:** Distribution of time spent consecutively seeking a PCR test before receiving one. The quadrant settings and color/hash schemes in **C** and **D** are identical to those in **B**.

In contrast to Scenario A, we find that test capacity *N* = 300 is met essentially every day for each quadrant (Figure 2**C**). Despite this uniformity, temporal crowding in Figures 2**D**(i) and 2**D**(iii) is less severe compared to Figures 2**D**(ii) and 2**D**(iv). This is because in Figures 2**D**(i) and 2**D**(iii), during each simulation, supply crowding is generally severe when COVID-19 infections are high (i.e. many test-seeking agents do not receive a test on a given day), but relatively mild otherwise (see Figure S5). Meanwhile, in Figures 2**D**(ii) and 2**D**(iv), due to high *P*_ILI_, supply crowding is essentially always severe throughout the duration of each simulation (Figure S5). Despite this, the trend of worsening test timeliness with decreasing LOD (Figure 2**A**) is still primarily due to the detection of residual viral load, as seen by the broadening of the overall curves in Figures 2**B**(iii) and 2**B**(iv). The waiting time distributions corresponding to the “COVID-19 only” infection status are plotted in Figure S6, and as can be seen, the distributions at low and high ILI are similar (compare Figures S6(i) and S6(iii) against Figures S6(ii) and S6(iv)). This similarity arises because even for low ILI, agents with the “COVID-19 only” status are testing during the outbreak wave where supply crowding is of a similar order of severity to when ILI is high (Figure S5). However, the marginally higher supply crowding with high ILI (Figure S5) results in the waiting time distributions being slightly broader at high ILI compared to low ILI in Figure S6.

### 4.3 Scenario C

In Scenario C, we introduce RATs as a diagnostic tool for COVID-19 to complement PCR tests. However, we assume that RAT supply is limited, approximating a middle pandemic “era” where these tools become available but their distribution is restricted by the government due to supply limitations. In this scenario, eligibility for a RAT is stringent, requiring the agent to be symptomatic, and to obtain a PCR test, the agent must have received a positive RAT within the last 3 days.

One of the stated concerns with the wide-spread use of RATs in Quebec during the COVID-19 pandemic was the utility of this tool given potentially lower sensitivity and specificity rates [46]. Consequently, in this scenario a primary concern is to evaluate the timeliness of the RAT and PCR test signals as sensitivity/specificity varies. The sensitivity/specificity of a RAT is influenced by both its *LOD* and its probability of failing for reasons unrelated to *LOD*, such as a lack of adherence to the manufacturer’s instructions and recommended test conditions [47]. In this scenario, we focus on test failure probability as a driver of sensitivity/specificity and assume RATs are just sensitive enough to indicate infectiousness by setting *LOD* = 6 [14]. We additionally fix PCR *LOD* at 6 such that both RATs and PCR tests cannot detect residual viral load in agents who are no longer infectious with COVID-19. We then explore signal timeliness as a function of RAT *P*_fail_ and *P*_ILI_. Note that by setting RAT *LOD* to 6, RAT sensitivity and specificity are both equal to 1 − *P*_fail_.

Timeliness heatmaps are depicted in Figure 3**A** for RATs and Figure 3**B** for PCR tests. Note that outlier values of RAT signal timeliness for high RAT failure probabilities, indicated by squares shaded in red, arise due to the cross correlation function becoming unresolvable. This happens because at extremely high *P*_fail_ and in the presence of background ILI, all available RATs are consumed daily, resulting in a daily positive RAT count of ∼ *P*_fail_ × *N* ≈ 300, where *N* is the number of RATs available daily (see Figures S7 and S8).

**Fig 3.**
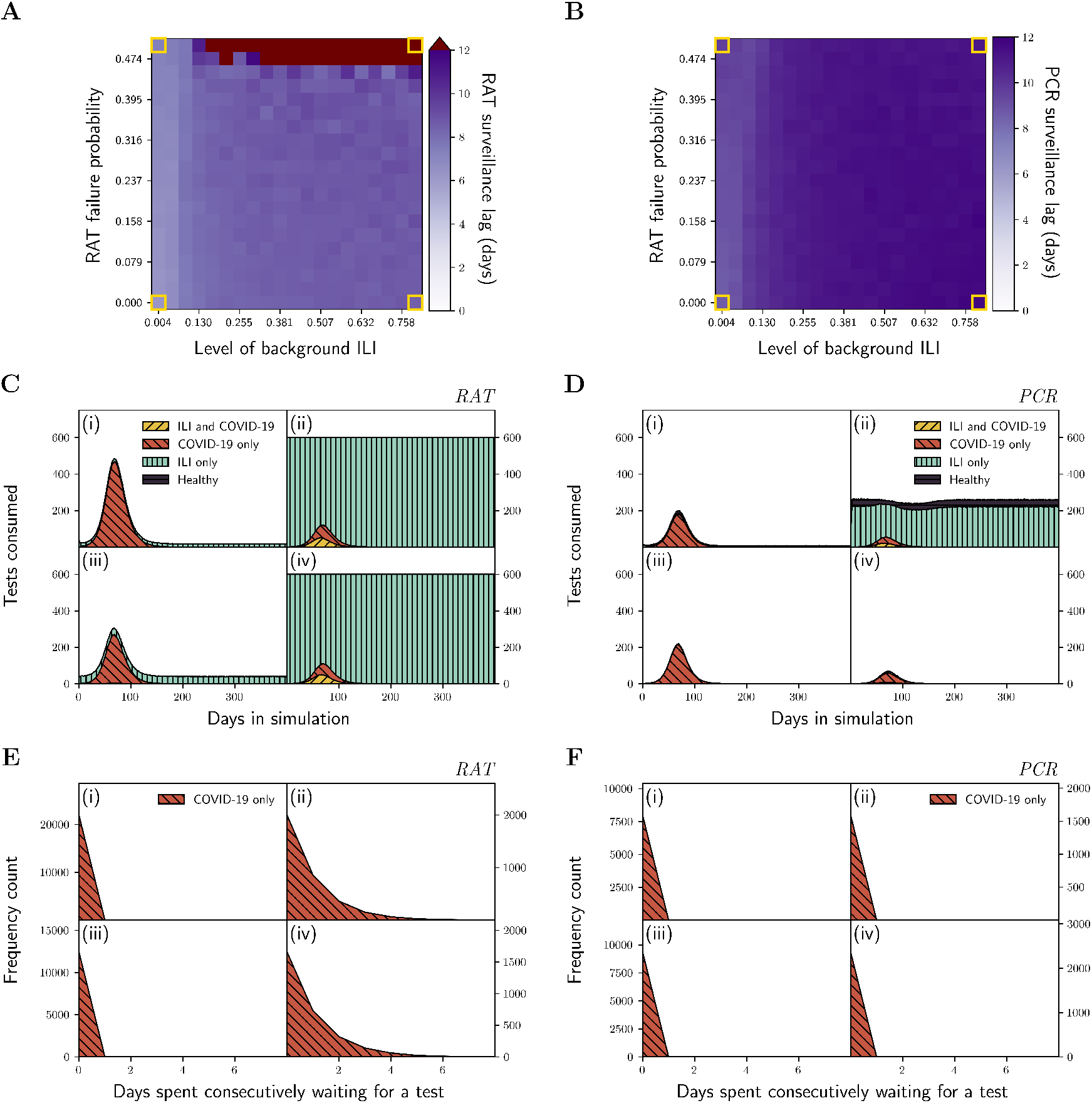
**A:** Surveillance timeliness of RATs in Scenario C as a function of *P*_ILI_ and RAT *P*_fail_. A positive timeliness value indicates that the surveillance signal (daily positive RATs) lags the pandemic signal (daily new infections). Yellow highlighted boxes at the heatmap corners identify settings of (*P*_ILI_, *P*_fail_) used to construct the corresponding quadrants in the **C** and **E** subplots. Red heatmap cells represent timeliness values that extend above 12 days, and comprise outliers which arise from an unresolvable cross correlation function (see Figures S7A(ii) and S8A(ii)). **B:** Surveillance timeliness of PCR tests in Scenario C as a function of *P*_ILI_ and RAT *P*_fail_. Yellow highlighted boxes at the heatmap corners identify settings of (*P*_ILI_, *P*_fail_) used to construct the corresponding quadrants in the **D** and **F** subplots. **C**: Daily number of RATs consumed. The quadrant settings are (i) (*P*_ILI_, *P*_fail_) = (0.004, 0.500), (ii) (*P*_ILI_, *P*_fail_) = (0.800, 0.500), (iii) (*P*_ILI_, *P*_fail_) = (0.004, 0.000), and (iv) (*P*_ILI_, *P*_fail_) = (0.800, 0.000). Furthermore, for each quadrant, the curve is partitioned by infection status at the time of testing, each associated with a unique color/hash combination. **D:** Daily number of PCR tests consumed. The quadrant settings and color/hash scheme are identical to **C. E:** Distribution of days spent consecutively seeking a RAT before receiving one. The quadrant settings are identical to **C**, but only the “COVID-19 only” infection status is shown. **F:** Distribution of days spent consecutively seeking a PCR test before receiving one. The quadrant settings are identical to **D**, but only the “COVID-19 only” infection status is shown.

In general, we observe that higher *P*_ILI_ leads to worse RAT surveillance timeliness, and interestingly there is only a mild dependence of RAT timeliness on *P*_fail_ (see Figure 3**A**). Since detection of residual viral load is no longer possible, this trend arises solely due to supply and temporal crowding. As seen in Figure 3**C**(ii) and 3**C**(iv), there is RAT supply crowding at higher *P*_ILI_ since RATs can no longer be allocated to all agents who request one and test consumption saturates at the RAT capacity of *N* = 600 daily. In contrast, test capacity is never reached at low *P*_ILI_, even during the COVID-19 infection peak. The upshot of supply crowding is that agents with COVID-19 experience temporal crowding, thus delaying the surveillance signal. To illustrate this, in Figure 3**E**, we plot the distribution of the number of days an agent infected with only COVID-19 spends seeking a RAT prior to acquiring one. As seen in Figure 3**E**, for a given RAT *P*_fail_, increased *P*_ILI_ leads to a greater proportion of agents spending more time waiting to receive a RAT (compare Figures 3**E** (i) and (ii) as well as Figures 3**E** (iii) and (iv). Note that in Figure 3**E**, while we focus on agents with only COVID-19 at the time of testing, we observe nearly identical trends for all categories of infection status (see Figure S9**A**).

The timeliness of PCR tests follows similar trends to that of RATs, although all timeliness values are higher due to the requirement of having a positive RAT result in order to obtain a PCR test. This requirement extends the time between being exposed to COVID-19 and receiving a positive test result for PCR tests compared to RATs (Figure 3**B**). Since the highest RAT failure probability we consider is *P*_fail_ = 0.5 and the daily PCR test capacity is taken to be *N* = 300, we do not observe supply crowding or temporal crowding (see Figures 3**D** and 3**F** respectively). The absence of temporal crowding in Figure 3**F** is also observed when all infection statuses are included (see Figure S9**B**). We note that between Figure 3**D** (i) and (iii), there is a similar number of agents who take a PCR test and only have COVID-19, even though Figure 3**D**(i) corresponds to a much larger RAT failure rate. This observation arises due to the fact that an agent with COVID-19 who receives a false negative RAT may test again the next day if they are still symptomatic. While a single RAT has a high probability of failing, thereby returning a negative result for agents infectious with COVID-19, several repeated tests are less likely to all fail. Therefore, the magnitude of agents who take a PCR test and only have COVID-19 is not substantially reduced in Figure 3**D**(i) compared to Figure 3**D**(iii). Note that in contrast, due to the repeated testing from false negatives, there are clearly more RATs consumed in Figure 3**C** (i) compared to (iii). The varied RAT consumption is not due to a difference in the number of COVID-19 infections, since they are nearly identical between Figure 3**C** (i) and (iii) (compare Figure S7A (i) and (iii)). We also note that a relatively large population of healthy agents emerges in Figure 3**D**(ii), which is mostly absent in Figure 3**D**(iv). To see why, note that in 3**D**(ii), RATs have a high probability of failing and there is a high level of background ILI. Consequently, a substantial number of agents with ILI receive a false positive result when they take a RAT, with many of these false positives occurring within the 3 days prior to ILI recovery. As a result, by the time they take a PCR test, the agent may have already recovered from their ILI. Since we prohibit back-to-back ILI infections of particular agents, recovery from ILI implies that an agent must subsequently have entered a healthy state or contracted COVID-19. When *P*_ILI_ is decreased in 3**D**(i), there are fewer ILI-related false positives, and as such a much smaller number of healthy agents receive PCR tests.

### 4.4 Scenario C^*^

In Scenario C^*^, we consider the same dual-test system as in Scenario C, except we increase RAT test capacity to *N* = 20000 such that the entire population can take a RAT on any given day. The corresponding timeliness heatmaps are shown in Figures 4 **A** and **B**.

**Fig 4.**
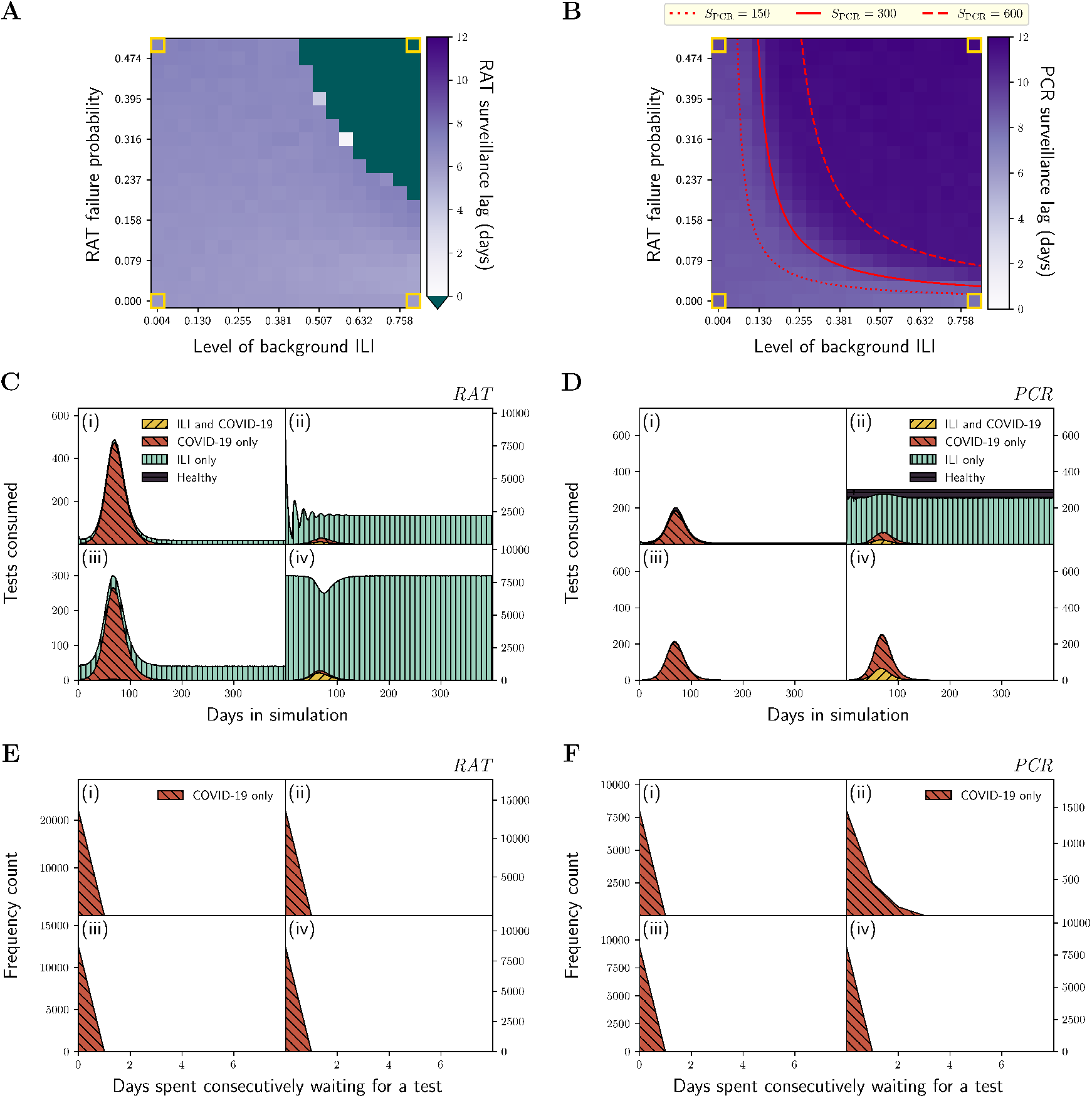
**A:** Surveillance timeliness of RATs in Scenario C^*^ as a function of *P*_ILI_ and RAT *P*_fail_. A positive timeliness value indicates that the surveillance signal (daily positive RATs) lags the pandemic signal (daily new infections). Yellow highlighted boxes at the heatmap corners identify settings of (*P*_ILI_, *P*_fail_) used to construct the corresponding quadrants in the **C** and **E** subplots. Green heatmap cells represent timeliness values that extend below 0 days, and comprise outliers which arise from an unresolvable cross correlation function (see Figures S10A(ii) and S11A(ii)). **B:** Surveillance timeliness of PCR tests in Scenario C^*^ as a function of *P*_ILI_ and RAT *P*_fail_. Yellow highlighted boxes at the heatmap corners identify settings of (*P*_ILI_, *P*_fail_) used to construct the corresponding quadrants in the **D** and **F** subplots. The red curves correspond to different values of *S*_PCR_ (dotted: *S*_PCR_ = 150, solid: *S*_PCR_ = 300, dashed: *S*_PCR_ = 600), which is the daily number of requested PCR tests at steady state in the compartmental approximation of the ABM (see the Supplementary Material and Figure S3). Each red curve defines the locus of points of (*P*_ILI_, *P*_fail_) which yield a value of *S*_PCR_ in the compartmental model. **C**: Daily number of RATs consumed. The quadrant settings are (i) (*P*_ILI_, *P*_fail_) = (0.004, 0.500), (ii) (*P*_ILI_, *P*_fail_) = (0.800, 0.500), (iii) (*P*_ILI_, *P*_fail_) = (0.004, 0.000), and (iv) (*P*_ILI_, *P*_fail_) = (0.800, 0.000). Furthermore, for each quadrant, the curve is partitioned by infection status at the time of testing, each associated with a unique color/hash combination. **D:** Daily number of PCR tests consumed. The quadrant settings and color/hash scheme are identical to **C. E:** Distribution of days spent consecutively seeking a RAT before receiving one. The quadrant settings are identical to **C**, but only the “COVID-19 only” infection status is shown. **F:** Distribution of days spent consecutively seeking a PCR test before receiving one. The quadrant settings are identical to **D**, but only the “COVID-19 only” infection status is shown.

As with Scenario C, high RAT consumption under certain conditions results in positive RAT curves without a clearly defined peak, generating uninterpretable cross-correlation results. In particular, at high *P*_fail_ and high *P*_ILI_, a large fraction of the symptomatic population who take a RAT test receive a positive RAT result on day 0, making them ineligible to test again for *T*_+_ = 14 days. Since these periods of re-test delay are initially synchronized, we observe oscillations in the number of positive tests received over time, confounding the ability to determine a cross-correlation with the infection curve (see Figures S10A(ii) and S11A(ii)). The conditions corresponding to this situation are shaded in green in Figure 4**A**. Additionally, supply crowding of RATs no longer exists in Scenario C^*^, since any agent who requests a RAT will receive one (see Figure 4**C**). Consequently, at high *P*_ILI_, agents do not see increased test waiting times for RATs, so temporal crowding also disappears (Figure 4**E**).

With an unlimited supply of RATs, PCR surveillance timeliness exhibits a clear dependence on *P*_fail_ and *P*_ILI_, whereby timeliness appears to worsen along a critical boundary in Figure 4**B**. Like Scenario C, these variations in timeliness are connected to temporal crowding, which itself is caused by supply crowding. Evidently, we see that in Figure 4**D**, supply crowding only exists in quadrant (ii) when *P*_ILI_ and *P*_fail_ are both large, such that many agents are (1) seeking a RAT for a reason other than having COVID-19 (Figure 4**C**(ii)), and (2) receiving a false positive. Correspondingly, in Figure 4**F** we only observe temporal crowding, i.e. long tails in the waiting time distributions, in quadrant (ii). Like Scenario C, while we focus on agents with only COVID-19 at the time of testing in Figures 4**E** and 4**F**, we observe nearly identical trends when all categories of infection status are included (see Figure S12).

To obtain a more precise understanding of how supply crowding influences PCR timeliness in Scenario C^*^, we seek to determine an approximate analytic expression for the daily number of PCR tests requested at steady state in the absence of COVID-19 infections, *S*_PCR_, as a function of *P*_fail_ and *P*_ILI_. We derive Equation 7 by constructing a compartmental model which approximates the dynamics of our ABM with ILI circulation only, with details provided in the Supplementary Material. Essentially, we seek values of *P*_fail_ and *P*_ILI_ for which PCR test capacity is already saturated even in the absence of any COVID-19 infections. Solving our compartmental model at steady state gives:

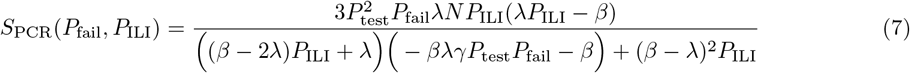

In Equation 7, λ = 8.5 days is the mean time to recovery from an ILI infection, *β* := *T*_+_ = 14 days is the time restricted from testing since taking a will-be-positive RAT, *N* = 20000 is the population size, *P*_test_ = 0.5 is the probability of seeking a test for eligible agents, and *γ* = 1/day is the rate that eligible agents are allowed to take tests. The level curve formed by setting *S*_PCR_ = 300, shown as a solid red line in Figure 4**B**, tightly corresponds to the main curvature of the PCR surveillance timeliness heatmap. Additional level curves for different levels of *S*_PCR_ (*S*_PCR_ = 150 and *S*_PCR_ = 600) are also shown for reference in Figure 4**B**. As *S*_PCR_ increases, the level curve shifts towards the upper right, which suggests that surveillance systems with larger PCR test capacities would tolerate greater RAT *P*_fail_ and *P*_ILI_ before timeliness worsens.

Ultimately, Equation 7 provides a way to estimate whether the PCR test signal is timely for a given extent of ILI circulation, assuming known values of the daily PCR test capacity and the failure rate of RATs when they are available in unlimited supply. We note that the analytic expression in Equation (7) is an approximation only as it omits several subtleties captured by the ABM including simultaneous agent transitions between two-states (see the discussion in the Supplementary Material) and “higher-order” or more rapid additional transition pathways between states. Unlike the ABM, Equation (7) also omits the exclusion from PCR testing of agents who took a PCR test in the previous *T*_*−*_ = 14 days that returned a true negative, or a PCR test in the previous *T*_+_ = 90 days that returned a false positive.

## 5 Discussion

Our findings demonstrate that the timeliness of test-based surveillance systems can evolve in important ways over the course of a pandemic depending on the epidemiological context as well as the quality and quantity of the test in question. In Scenarios A and B, we considered conditions representative of early stages of the COVID-19 pandemic, where only PCR tests were available. Here, we found that the high sensitivity of the PCR tests, and their ability to detect residual viral loads in recovered patients, strongly impacted test timeliness. When testing criteria was extremely stringent such that only symptomatic individuals could receive a PCR test (Scenario A), this high test sensitivity did not compromise surveillance timeliness for low background ILI, since only agents actively infected with COVID-19 were seeking tests. However, for high background ILI, these highly sensitive tests detected residual viral loads in agents with ongoing ILI infections but who were not infectious with COVID-19, compromising surveillance timeliness. When test eligibility criteria was broadened (Scenario B), surveillance timeliness was no longer sensitive to background ILI level, since healthy agents could also receive tests. However, the detection of residual viral load still led to a dependence of the surveillance timeliness on PCR test LOD.

We next considered scenarios intended to reflect later stages of a pandemic, when home-based RATs are also available, first in limited (Scenario C) then in unlimited (Scenario C^*^) supply. We considered a wide range of probabilities for RAT failure (i.e. the return of the wrong test result), in line with original concerns regarding their widespread use. In Scenario C, we found that the high demand for RATs at high levels of background ILI led to supply crowding due to limited test availability as well as temporal crowding as agents had to wait longer to receive a RAT. Critically, this included the subset of agents with the “COVID-19 only” infectious status (at the time of testing), generating a surveillance lag. Additionally, in this scenario with strict symptom-based testing requirements for RATs, surveillance timeliness is also relatively insensitive to the RAT failure probability. This is because the daily number of new positive RATs due to ILI demand is essentially a flat line whose magnitude increases with increasing *P*_fail_, while the timing of the additional demand peak from COVID-19 infections is minimally affected by *P*_fail_. The exception is when the RAT failure probability gets very high, and the cross-correlation calculation is corrupted. In Scenario C^*^, freeing the restriction on RAT capacity eliminates RAT supply/temporal crowding. However, when RAT failure probability and the level of background ILI are sufficiently high, PCR test crowding occurs, compromising the timeliness of this metric. In particular, although the RAT signal timeliness is relatively insensitive to the failure probability of the RATs, PCR surveillance becomes less tolerant to RAT failure probability when the level of background ILI increases. Indeed, at very high levels of background ILI, even small RAT failure probabilities begin to worsen the PCR signal timeliness, as also demonstrated by the steady-state analytic result based on an approximate compartmental model (the red lines in Figure 4**B** and Equation (7)). Therefore, Scenarios C and C^*^ demonstrate that while increasing RAT capacity can mitigate competition for RATs, downstream competition for PCR tests may limit the utility of the PCR test signal as a surveillance tool if RAT tests are not sufficiently high quality and other symptom-causing pathogens are in circulation.

## 6 Caveats

Our model makes several key simplifying assumptions. First, we assume several variables to be constant in time, when in reality they vary. For example, we have mentioned that ILI fluctuates seasonally, but the rate of ILI is fixed in our simulations. We have also taken the probability of seeking a COVID-19 test to be constant, yet test seeking behaviour is known to be time-varying (e.g. for influenza [48]). Indeed, during the COVID-19 pandemic, test seeking was associated with time-varying factors such as disease susceptibility (e.g., vaccination status), the state of public health measures including NPIs (e.g., lockdowns), and risk perception (which may in turn modulate an individual’s adherence to NPIs) [49, 50]. Second, our model assumes homogeneity among agents in terms of ILI prevalence and test-seeking behavior, while in reality these factors may be influenced by factors such as age and geographical location. Note that some spatial heterogeneity is inherently modeled in our simulations with Synthpops, as contact patterns in Synthpops populations depend on the population “layer” in which contacts occur (e.g., community, school, work, and household). Future work can attempt to generalize this framework to refine our understanding of population heterogeneities on surveillance system timeliness. Third, to measure how timeliness is impacted by the evolution of the test signal only, we do not incorporate behavioural changes in response to test results, such as isolation or quarantine. In reality, the occurrence of these responses would alter the infection signal, with important consequences for test timeliness. Finally, we do not consider pathogen-pathogen interactions in our simulations or the implementation of NPIs such as mask-wearing or social distancing, which may both affect patterns of transmission and susceptibility.

## 7 Conclusion

Our findings highlight important considerations when designing test-based surveillance systems. First, we show that surveillance system parameters can influence surveillance timeliness through mechanisms that may not be initially obvious. For example, Scenarios A and B demonstrate how highly sensitive tests, which are accurate from a diagnostic perspective, may worsen surveillance timeliness by detecting residual viral load. Meanwhile, Scenarios C and C^*^ highlight how modifying the quality and quantity of RATs can generate testing bottlenecks that lag the downstream PCR surveillance signal when background levels of co-circulating, symptom-causing illnesses are substantial. Tests vary not only in terms of their diagnostic sensitivity and specificity, but also in terms of their cost and user-friendliness. Therefore, public health and socioeconomic objectives should be carefully weighed to guide the choice of surveillance systems adopted depending on the current epidemiological context and the quality and quantity of available testing and diagnostic technologies.

## Data Availability

All data and code needed to reproduce the results will be made publicly available on Zenodo upon acceptance. An interactive tool which allows for a detailed exploration of how the underlying simulation dynamics change in response to key parameters will also be made available at this time.

https://github.com/synthpops/synthpops/blob/main/synthpops/data/usa-Washington-seattle_metro.json

https://data-explorer.oecd.org/vis?pg=0&bp=true&snb=10&tm=enrolment%2520rates&vw=tb&df%5Bds%5D=dsDisseminateFinalDMZ&df%5Bid%5D=DSD_EAG_UOE_NON_FIN_STUD%2540DF_UOE_NF_ENRL_RATE&df%5Bag%5D=OECD.EDU.IMEP&df%5Bvs%5D=1.0&dq=GBR._T......A......_T.Y_GE65%252BY60T64%252BY55T59%252BY50T54%252BY49%252BY48%252BY47%252BY46%252BY45%252BY44%252BY43%252BY42%252BY41%252BY40%252BY39%252BY38%252BY37%252BY36%252BY35%252BY34%252BY32%252BY33%252BY31%252BY30%252BY29%252BY28%252BY27%252BY26%252BY24%252BY25%252BY23%252BY22%252BY21%252BY20%252BY19%252BY18%252BY17%252BY16%252BY15%252BY14%252BY13%252BY12%252BY11%252BY10%252BY9%252BY8%252BY7%252BY6%252BY5%252BY4%252BY3&pd=2022%252C2022&to%5BTIME_PERIOD%5D=false&isAvailabilityDisabled=false

https://www.ons.gov.uk/peoplepopulationandcommunity/birthsdeathsandmarriages/families/datasets/familiesandhouseholdsfamiliesandhouseholds/current

https://www.ons.gov.uk/employmentandlabourmarket/peopleinwork/employmentandemployeetypes/datasets/employmentunemploymentandeconomicinactivitybyagegroupseasonallyadjusteda05sa/current

https://www.ons.gov.uk/businessindustryandtrade/business/activitysizeandlocation/datasets/ukbusinessactivitysizeandlocation

## Author contributions

R.K.A, R.Y., N.D., and C.E.W. conceived of the idea. R.Y., P.N.N.T., A.S.-J., J.Y.L., and N.D. developed the software and documentation. R.Y. performed the simulations, and R.Y. and C.E.W. performed the analyses. R.Y. and C.E.W. wrote the paper. All authors reviewed and edited the manuscript.

