## Supporting Information for "Evaluating the evolution of the timeliness of test-based surveillance systems over the course of a pandemic"

**This PDF file includes:**

Supporting text
Figs. S1 to S12
Table S1
SI References

### Supporting Information Text

**Population construction using a variation of SynthPops.** For our experiments, we generate a fixed population parameterized primarily by United Kingdom (UK) demographic data using a custom variation of SynthPops. We retrieve binned population age distribution data from UK.GOV which was initially recorded in a 2011 census (1). We assume an individual's age is at most 100, and to accommodate this assumption of an upper age limit, we approximate the proportion of individuals belonging to the age bin $[75, 100]$ as the proportion of individuals aged 75+ reported in the UK.GOV data. Employment rates by age are taken from the UK Office for National Statistics (ONS), and we calculate employment rates for each age bin using a monthly average of data from January 2021 to December 2021, which was released on April 12, 2022 (2). When an employment rate is specific to an age *range*, we apply the same employment rate to each age in the range. For the employment rate of individuals belonging to the age bin $[65, 100]$, we rely on the 2021 average employment rate of individuals aged 65+ reported in the UK ONS data. We set school enrollment rates by age using data from the Organisation for Economic Co-operation and Development (OECD), specific to the UK in 2022 (3). We assume individuals with ages between 0 and 2 (inclusive) do not attend school. When an enrollment rate is specific to an age *range*, we apply the same enrollment rate to each age in the range. For the enrollment rate of individuals belonging to the age bin $[65, 100]$, we use the enrollment rate of individuals aged 65+ reported in the OECD data. We retrieve household size distribution data from the UK ONS and use estimates from 2022, taken from data released on May 8, 2024 (4). We assume household size is at most 7 and we normalize the household size distribution such that it sums to 1. For the proportion of households with size 7, we use the proportion of households of size 7+ in the UK ONS data. We set the distribution of workplace size, in terms of number of personnel, using a 2022 edition of UK ONS data for enterprise sizes in the UK (5). We limit workplace size by interpreting the proportion of workplaces of size 250+ as the proportion of workplaces with size between 250 and 1999, inclusive. For the following population fields, due to data unavailability, we made approximations using existing United States (US) data supplied by SynthPops. First, we use data based on Seattle, Washington for the school size distribution by school type (e.g., elementary school, middle school, etc.) as well as the school type that different age bins belong to (e.g., elementary school for age 6 to 10 inclusive). Second, for the household head age distribution by family size, we take the general US household head age distribution for a family size of 3 and repeat it across all family sizes (from 1 to 7) using the same household head age brackets supplied in the general US data. In terms of generating a SynthPops population, our variation follows the SynthPops method but restructures the code for greater modularity. The generated population retains a population structure with household, school, workplace, and community layers, and we disable the optional long-term care facility layer. Note that we enable the differentiation of schools by type, which is an option that can be toggled in SynthPops.

**Model of background influenza-like-illness.** Let $s$ be the population size. We assume that a constant proportion $P_{\mathrm{ILI}}$ of the original population size (discounting any deaths) is infected with ILI throughout a simulation. Agents who die in the simulation due to COVID-19 cannot be infected with ILI. We assume that agents with ILI recover within $t \sim \mathcal{U}(7, 10)$ days, and we assume they are symptomatic for the entire duration of illness. Now suppose on day 0 of a simulation we select $N_{\mathrm{infect}} := \lfloor P_{\mathrm{ILI}} \cdot s \rfloor$ agents to be infected with ILI. Then, the first batch of ILI recoveries will be constrained between days 7 and 10. This means that no recoveries will occur prior to day 7, as illustrated in Figure S1(i). Instead, to simulate ILI that has been co-circulating for a prolonged period of time, we apply the following procedure. First, on day 0 of the simulation, we randomly select $N_{\mathrm{infect}}$ agents to be infected with ILI, and we initialize $N_{\mathrm{infect}}$ many recovery dates $d \sim \mathcal{U}(7, 10)$ to be transformed. Then, iterating over 1000 hypothetical days, whenever a recovery date $d$ is reached, it is replaced with a new recovery date $d + t$ where $t \sim \mathcal{U}(7, 10)$, to simulate the replacement of a recovered agent with a new agent infected with ILI. This procedure outputs a recovery day distribution and we shift the earliest recovery date to day 1 (see sample distributions in Figure S1(ii)). The distribution transformed in this manner is then assigned as the recovery day distribution for the $N_{\mathrm{infect}}$ agents selected to have ILI on day 0 of the actual simulation.

**Model of viral load.** In Figure S2 we depict sample viral load trajectories computed with Covasim (Figure S2(i)) and with our custom method (Figure S2(ii)). Note that if an agent is infectious on a given day, then their relative transmissibility is defined by Covasim in Equation 1 with time-dependent viral load $V$ as a multiplicative factor. If an agent is not infectious on a given day, then their relative transmissibility is 0.

$$T := T' \times f_{\mathrm{quar}} \times f_{\mathrm{asymp}} \times f_{\mathrm{iso}} \times \beta_{\mathrm{layer}} \times V \qquad [1]$$

Here, $T'$ is a baseline value of the agent's relative transmissibility to be updated by Equation 1, $f_{\mathrm{quar}}$ is the agent's quarantine factor, $f_{\mathrm{asymp}}$ is the agent's asymptomatic factor, $f_{\mathrm{iso}}$ is the agent's isolation factor, and $\beta_{\mathrm{layer}}$ is a factor associated with the setting/layer in which disease transmissions are being computed. Due to the integral role of $V$ in Equation 1, we rescale our custom viral load trajectories to preserve the general transmission dynamics of Covasim (see Figure S2(iii)). We perform this rescaling as follows. Let $a = 0.75$, $b = 2.00$, and suppose an agent has viral load $V$ computed with our custom method. Then $V_{\mathrm{rescaled}}$ is set to

$$V_{\mathrm{rescaled}} := a + (b - a) \times \frac{V - 6}{11 - 6} \qquad [2]$$

This rescaled value $V_{\mathrm{rescaled}}$ is then substituted for $V$ in Equation 1 to calculate the agent's relative transmissibility.

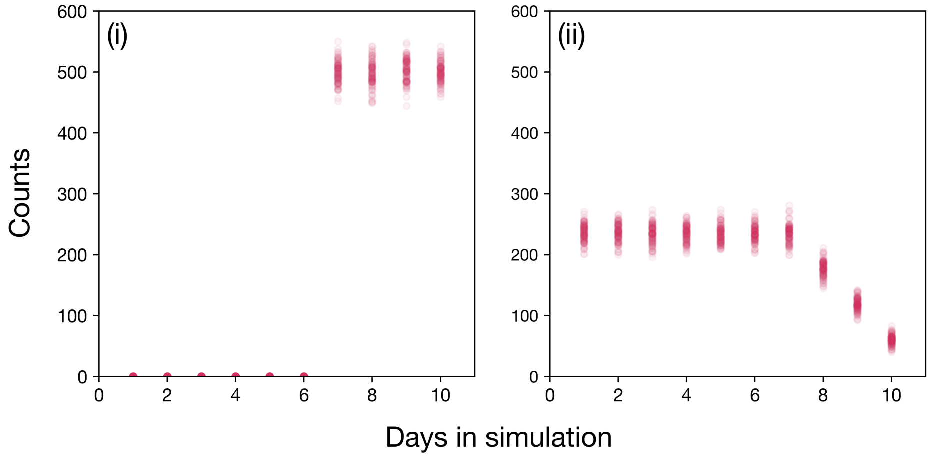


**Fig. S1.** 100 sample distributions of ILI recovery dates for a population size of $s = 20000$ with a 10% rate of ILI. (i) Recovery day distributions with each obtained by instantiating $N_{\text{infect}}$ infections and sampling $N_{\text{infect}}$ recovery dates $d \sim \mathcal{U}(7, 10)$. (ii) Recovery day distributions with each obtained by transforming an initial distribution (where $N_{\text{infect}}$ dates are sampled from $\mathcal{U}(7, 10)$) using the described method over 1000 days. The 1000 days hypothetically occur prior to the start of the actual simulation.

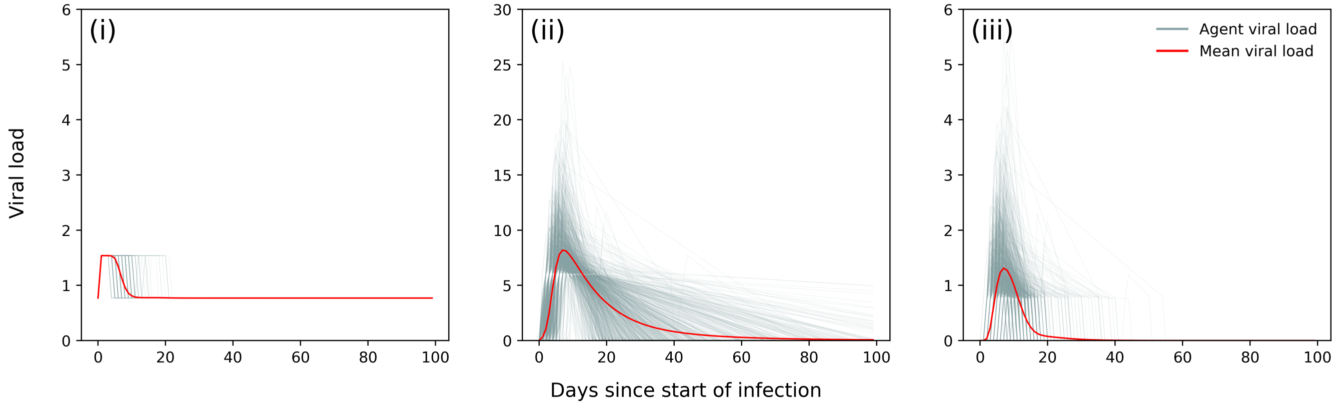


**Fig. S2.** Sample viral load trajectories produced by Covasim's kinetics model and our model for 1000 agents. (i) Viral load trajectories provided natively by Covasim. (ii) Viral load trajectories computed using our custom method described in the main text. (iii) Viral load trajectories computed with our method with additional rescaling.

**Compartmental approximation used for Scenario C$^*$.** To develop an approximate compartmental model, we make the following key assumptions:

1. No agents in the population have COVID-19: only ILI is in circulation in the population. This can either be interpreted as the situation before COVID-19 is introduced, or a long-term state after a COVID-19 epidemic has concluded.

2. Residual viral load is strictly below $10^6$ cp/ml in every agent such that any positive test always represents a false positive.

3. Deaths occur rarely enough such that the population size can be approximated to be a constant.

4. The state transition diagram follows that shown in Figure S3. Agents flow between being infected with ILI and eligible for RAT testing ($I_E$), healthy and eligible for RAT testing ($H_E$), infected with ILI and restricted from RAT testing ($I_R$), and healthy and restricted from RAT testing ($H_R$). We further split the $I_R$ compartment into $I_{HR}$ and $I_{IR}$ to denote agents infected with ILI and restricted from testing who were previously healthy and restricted from testing ($I_{HR}$ - i.e. these agents became infected) or infected with ILI and eligible for testing ($I_{IR}$ - i.e. these agents took a test and received a positive result). We do this because the transition flows available to these two groups of agents fundamentally differ, and so separate equations are required to determine the time rate of change of agents in these categories. Note that we disallow diagonal flows (i.e. $H_E \longleftrightarrow I_R$ and $H_R \longleftrightarrow I_E$) since they represent two-state changes (a change in disease state and in test restriction status). While these transitions are possible in the ABM framework, they are not obvious to model using the compartmental approach and happen relatively infrequently. We also specifically disallow the one-state transition $H_E \to H_R$ because a healthy agent cannot receive a RAT under Scenario C$^*$, and therefore will never become restricted from testing without first transitioning into an infected compartment. The complete list of variables for the compartmental model is shown in Table S1.

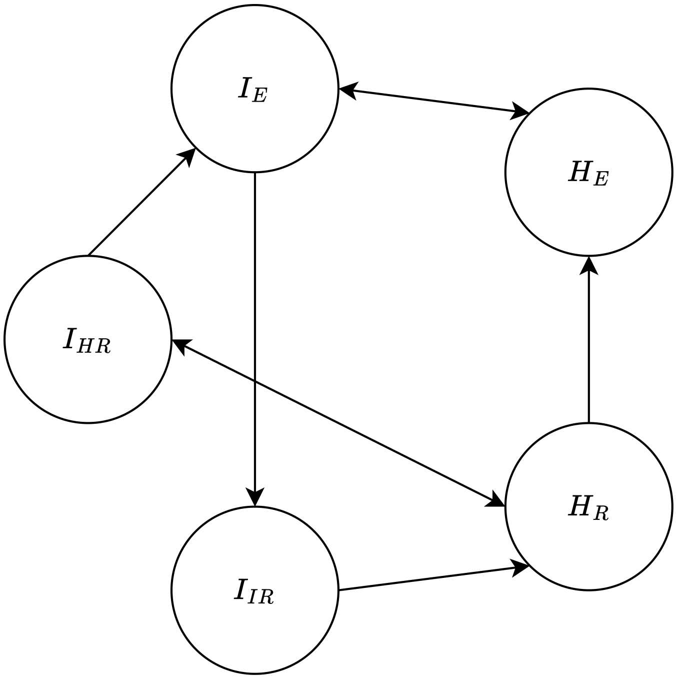


**Fig. S3.** State diagram summarizing the allowed "flows" between compartments.

| **Variable** | **Description** |
|---|---|
| $N = 20000$ agents | Population size |
| $\lambda = 8.5$ days | Mean time infected with ILI |
| $\beta = 14$ days | Time restricted from testing after taking a will-be positive RAT |
| $P_{\text{test}} = 0.5$ | Probability of eligible agents seeking a test |
| $\gamma = 1$/day | Rate that eligible agents are allowed to take tests |
| $P_{\text{fail}}$ | Probability of test failure |
| $P_{\text{ILI}}$ | Daily prevalence of ILI |
| $I_o$ | Total daily agents with ILI |
| $H_o$ | Total daily healthy agents |
| $I_E$ | Total daily ILI agents not restricted from testing |
| $I_{IR}$ | Total daily ILI agents restricted from testing whose previous state was "$I_E$" |
| $I_{HR}$ | Total daily ILI agents restricted from testing whose previous state was "$H_R$" |
| $H_E$ | Total daily healthy agents not restricted from testing |
| $H_R$ | Total daily healthy agents restricted from testing |

**Table S1.** Notation for variables used to construct compartmental model equations.

Given the above assumptions, we can then construct the following flow equations which govern the compartmental model:

$$\frac{dI_E}{dt} = -\gamma P_{\text{test}} P_{\text{fail}} I_E - \frac{1}{\lambda} I_E + \frac{I_o}{\lambda H_o} H_E + \frac{1}{\beta - \lambda} I_{HR} \quad [3]$$

$$\frac{dI_{IR}}{dt} = \gamma P_{\text{test}} P_{\text{fail}} I_E - \frac{1}{\lambda} I_{IR} \quad [4]$$

$$\frac{dI_{HR}}{dt} = \frac{I_o}{\lambda H_o} H_R - \frac{1}{\beta - \lambda} I_{HR} - \frac{1}{\lambda} I_{HR} \quad [5]$$

$$\frac{dH_E}{dt} = \frac{1}{\lambda} I_E - \frac{I_o}{\lambda H_o} H_E + \frac{1}{\beta - \lambda} H_R \quad [6]$$

$$\frac{dH_R}{dt} = \frac{1}{\lambda} I_{IR} + \frac{1}{\lambda} I_{HR} - \frac{I_o}{\lambda H_o} H_R - \frac{1}{\beta - \lambda} H_R \quad [7]$$

The following constraints must be satisfied at all times:

$$H_o = H_E + H_R = (1 - P_{\text{ILI}}) \times N \quad [8]$$

$$I_o = I_E + I_{IR} + I_{HR} = P_{\text{ILI}} \times N \quad [9]$$

Agents infected with ILI and eligible for testing ($I_E$) become ineligible for testing (flow to $I_{IR}$) at a daily rate $\gamma P_{\text{test}} P_{\text{fail}}$, i.e. the daily rate of seeking a test and having it fail into a false positive. In the ABM, these agents are restricted from retesting for

$\beta = 14$ days. Since $\beta > \lambda$, we do not permit agents to flow from $I_{IR}$ to $I_E$ in the compartmental model, since this would not be observed in the ABM. In practice, however, this flow would typically be included. Agents recover from all $I$ compartments at rate $\frac{1}{\lambda}$, transitioning to $H_E$ if they were in $I_E$ or $H_R$ if they were in $I_{IR}$ or $I_{HR}$. To maintain the proportion of agents with ILI constant, this same net rate of agent recovery (i.e. $\frac{1}{\lambda}(I_E + I_{IR} + I_{HR}) = \frac{I_o}{\lambda}$) must balance the rate that healthy agents become infected with ILI. Since agents are randomly selected from the healthy population for ILI infection, the fraction of agents becoming infected from $H_E$ is $\frac{H_E}{H_o}$ (and thus moving to $I_E$), and from $H_R$ is $\frac{H_R}{H_o}$ (and thus moving to $I_{HR}$). It is possible for agents to become infected with ILI in the ABM while still being excluded from testing from a prior ILI infection. In this case, their exclusion period will end in sooner than $\beta$ days, since they necessarily have already been excluded from testing for at least $\lambda$ days, i.e. the time it took them to recover. In reality, their exclusion period is possibly shorter than $\beta - \lambda$ since this does not account for the time they spent in the $H_R$ compartment. Nevertheless, we make the assumption that if they are infected with ILI while excluded from testing, the $H_R \longrightarrow I_{HR}$ transition happens very quickly, such that agents would become eligible for testing after $\beta - \lambda$ days. For simplicity, we ignore all other possibilities for state transitions.

At steady state, we can write the following system of three equations, involving equations 3, 4, and 6. Note that constraint equation 8 has been used to replace $H_R$, and constraint equation 9 has been used to replace $I_{HR}$.

$$0 = \underbrace{-\left(\gamma P_{\text{test}} P_{\text{fail}} + \frac{1}{\lambda} + \frac{1}{\beta - \lambda}\right)}_{A_1} I_E + \underbrace{\frac{I_o}{H_o \lambda}}_{B_1} H_E + \underbrace{\left(\frac{-1}{\beta - \lambda}\right)}_{C_1} I_{IR} + \underbrace{\frac{I_o}{\beta - \lambda}}_{D_1} \tag{10}$$

$$0 = \underbrace{\left(\gamma P_{\text{test}} P_{\text{fail}}\right)}_{A_2} I_E + \underbrace{\left(-\frac{1}{\lambda}\right)}_{C_2} I_{IR} \tag{11}$$

$$0 = \underbrace{\left(\frac{1}{\lambda}\right)}_{A_3} I_E + \underbrace{\left(-\frac{I_o}{H_o \lambda} - \frac{1}{\beta - \lambda}\right)}_{B_3} H_E + \underbrace{\frac{H_o}{\beta - \lambda}}_{D_3} \tag{12}$$

Solving the above linear system gives the following relationships:

$$H_E = -\frac{1}{B_3}(A_3 I_E + D_3) \tag{13}$$

$$I_{IR} = -\frac{A_2}{C_2} I_E \tag{14}$$

Substituting equations 13 and 14 into equation 10 then gives

$$\left(A_1 - \frac{B_1 A_3}{B_3} - \frac{C_1 A_2}{C_2}\right) I_E + \left(D_1 - \frac{B_1 D_3}{B_3}\right) = 0, \tag{15}$$

or

$$I_E = \frac{\frac{B_1 D_3}{B_3} - D_1}{A_1 - \frac{B_1 A_3}{B_3} - \frac{C_1 A_2}{C_2}}.$$

Re-introducing the definitions of $A_1, A_2, A_3, B_1, B_3, C_1, C_2, D_1, D_3$ as well as equations 8 and 9 we obtain

$$I_E = \frac{\frac{-I_o H_o}{(\beta-\lambda) I_o + \lambda H_o} - \frac{I_o}{\beta - \lambda}}{\frac{-\beta \gamma P_{\text{test}} P_{\text{fail}}}{\beta - \lambda} - \frac{\beta}{\lambda(\beta - \lambda)} + \frac{(\beta - \lambda) I_o}{\lambda((\beta-\lambda) I_o + \lambda H_o)}} \tag{16}$$

$$= \frac{-\frac{\lambda(\beta-\lambda) I_o H_o}{(\beta-\lambda) I_o + \lambda H_o} - \lambda I_o}{-\beta \lambda \gamma P_{\text{test}} P_{\text{fail}} - \beta + \frac{(\beta-\lambda)^2 I_o}{(\beta-\lambda) I_o + \lambda H_o}} \tag{17}$$

$$= \frac{\frac{-\lambda(\beta-\lambda) N P_{\text{ILI}}(1 - P_{\text{ILI}})}{(\beta - 2\lambda) P_{\text{ILI}} + \lambda} - \lambda N P_{\text{ILI}}}{-\beta \lambda \gamma P_{\text{test}} P_{\text{fail}} - \beta + \frac{(\beta-\lambda)^2 P_{\text{ILI}}}{(\beta - 2\lambda) P_{\text{ILI}} + \lambda}} \tag{18}$$

$$= \frac{\lambda N P_{\text{ILI}} (\lambda P_{\text{ILI}} - \beta)}{\Big((\beta - 2\lambda) P_{\text{ILI}} + \lambda\Big)\Big(-\beta \lambda \gamma P_{\text{test}} P_{\text{fail}} - \beta\Big) + (\beta - \lambda)^2 P_{\text{ILI}}} \tag{19}$$

Knowing $I_E$ is useful, because we can use it to directly estimate the daily number of PCR tests requested at steady state, denoted $S_{PCR}$. Given $I_E$ daily agents requesting a RAT at steady state, $P_{\text{test}} \times P_{\text{fail}} \times I_E$ agents will first seek a RAT, then receive one due to unlimited test capacity, and then have the test fail into a false positive. An agent is eligible for a PCR test if they received a positive RAT in the past 3 days, hence by accumulation, there will be an estimated $3 \times P_{\text{test}} \times P_{\text{fail}} \times I_E$

daily agents eligible for a PCR test at steady state. Finally, a proportion $P_{\text{test}}$ of such agents will actually seek a PCR test. Therefore, we find:

$$S_{PCR}(P_{\text{fail}}, P_{\text{ILI}}) = 3 \times P_{\text{test}}^2 \times P_{\text{fail}} \times I_E(P_{\text{fail}}, P_{\text{ILI}}), \quad [20]$$

or

$$S_{PCR}(P_{\text{fail}}, P_{\text{ILI}}) = \frac{3P_{\text{test}}^2 P_{\text{fail}} \lambda N P_{\text{ILI}}(\lambda P_{\text{ILI}} - \beta)}{\Big((\beta - 2\lambda)P_{\text{ILI}} + \lambda\Big)\Big(-\beta\lambda\gamma P_{\text{test}} P_{\text{fail}} - \beta\Big) + (\beta - \lambda)^2 P_{\text{ILI}}}. \quad [21]$$

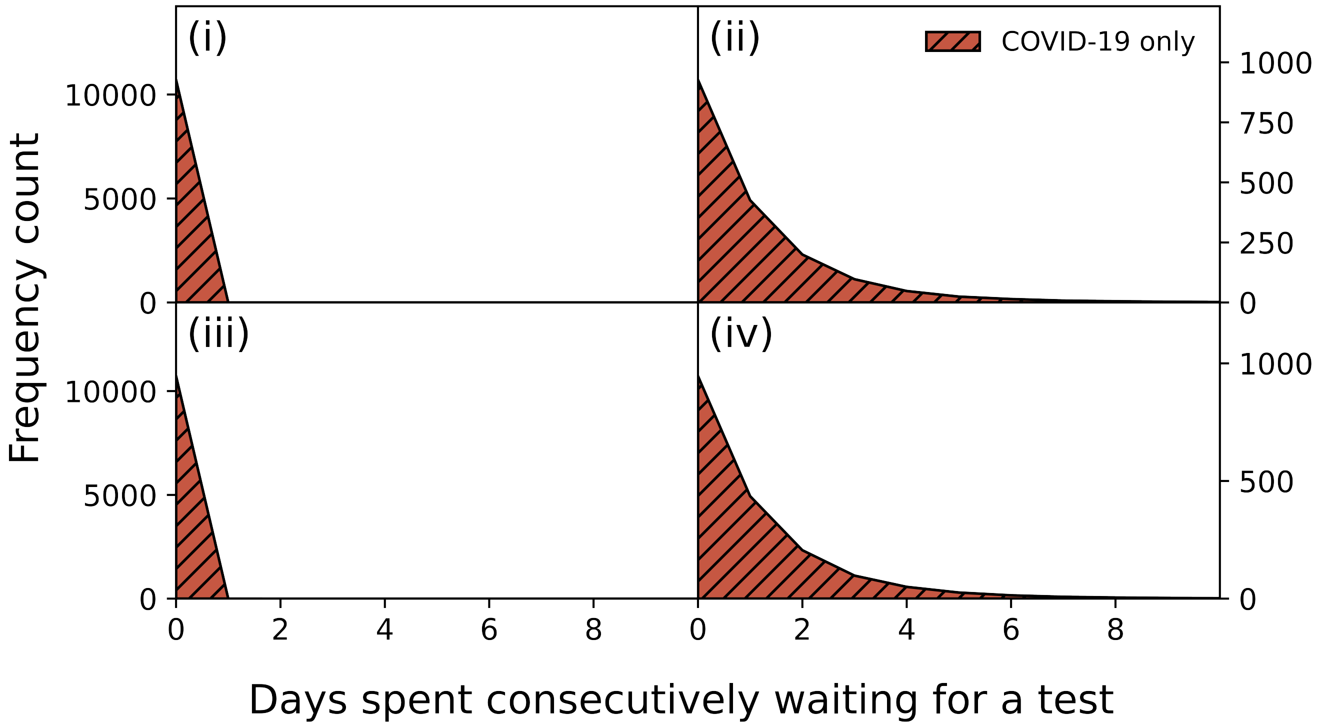


**Fig. S4.** Distribution of time spent consecutively seeking a PCR test before receiving one in Scenario A for agents who had only COVID-19 at the time of testing. The quadrant settings are (i) $(P_{\mathrm{ILI}}, LOD) = (0.004, 6)$, (ii) $(P_{\mathrm{ILI}}, LOD) = (0.800, 6)$, (iii) $(P_{\mathrm{ILI}}, LOD) = (0.004, 2)$, and (iv) $(P_{\mathrm{ILI}}, LOD) = (0.800, 2)$.

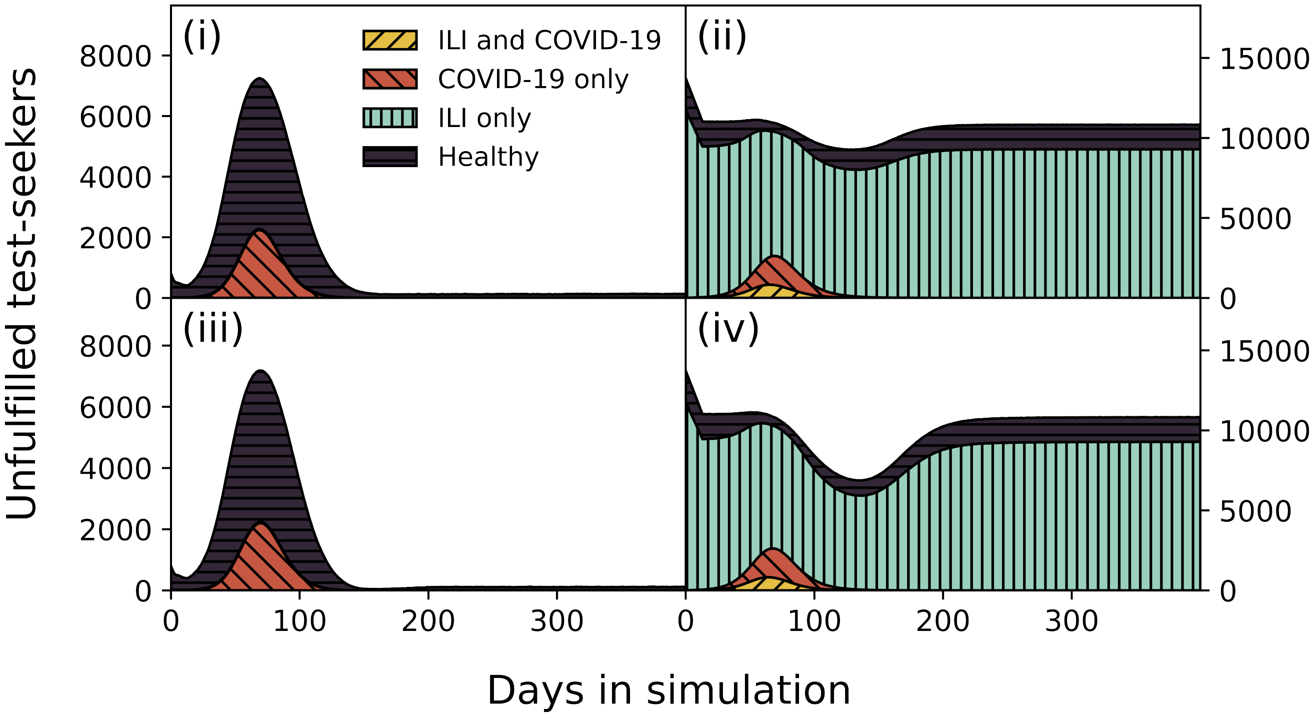


**Fig. S5.** Daily number of test-seeking agents who do not receive a test due to finite test capacity in Scenario B. The quadrant settings are (i) $(P_{\mathrm{ILI}}, LOD) = (0.004, 6)$, (ii) $(P_{\mathrm{ILI}}, LOD) = (0.800, 6)$, (iii) $(P_{\mathrm{ILI}}, LOD) = (0.004, 2)$, and (iv) $(P_{\mathrm{ILI}}, LOD) = (0.800, 2)$. Furthermore, for each quadrant, the curve is partitioned by infection status at the time of testing, each associated with a unique color/hash combination.

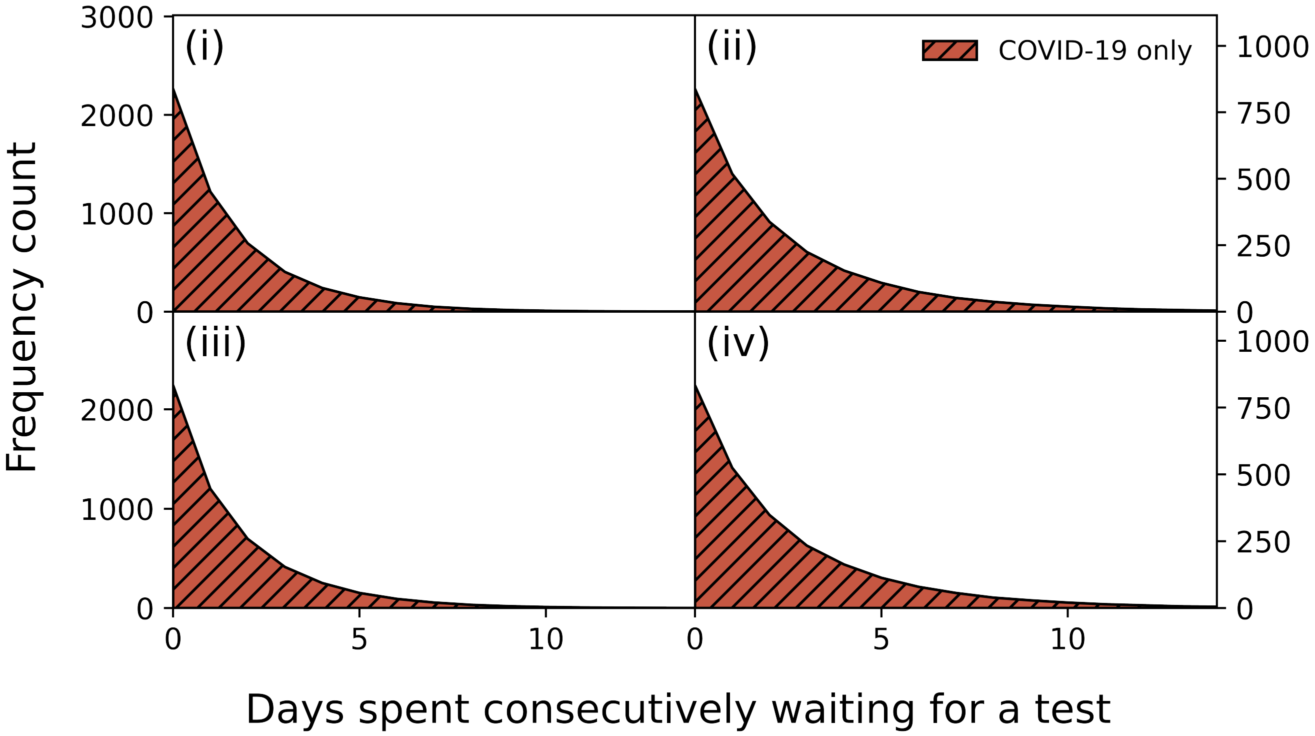


**Fig. S6.** Distribution of time spent consecutively seeking a PCR test before receiving one in Scenario B for agents who had only COVID-19 at the time of testing. The quadrant settings are (i) $(P_{\mathrm{ILI}}, LOD) = (0.004, 6)$, (ii) $(P_{\mathrm{ILI}}, LOD) = (0.800, 6)$, (iii) $(P_{\mathrm{ILI}}, LOD) = (0.004, 2)$, and (iv) $(P_{\mathrm{ILI}}, LOD) = (0.800, 2)$.

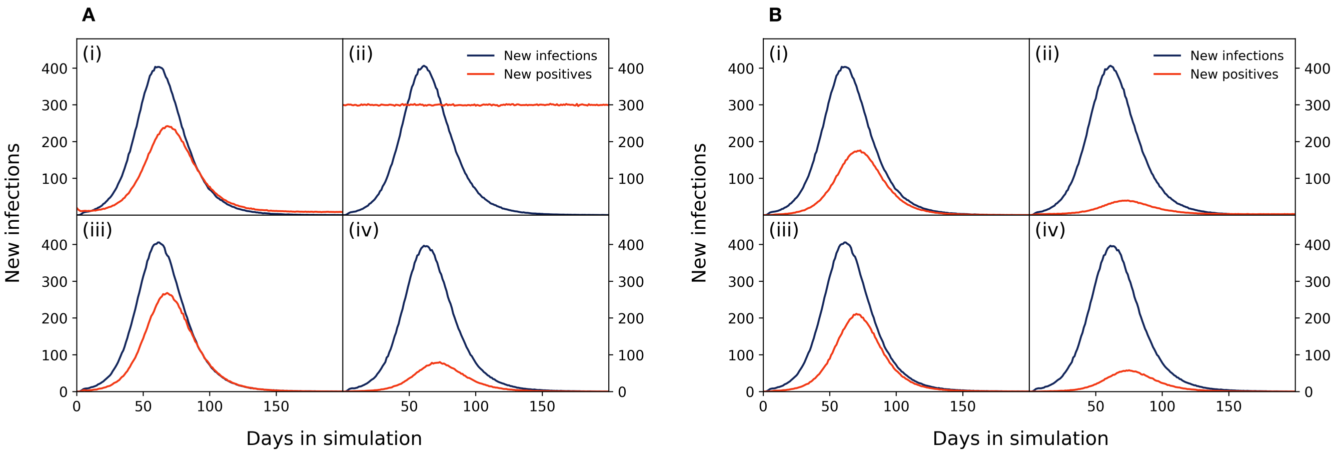


**Fig. S7.** **A:** Daily number of new infections and daily number of new positive RATs in Scenario C. The quadrant settings are (i) $(P_{\mathrm{ILI}}, P_{\mathrm{fail}}) = (0.004, 0.500)$, (ii) $(P_{\mathrm{ILI}}, P_{\mathrm{fail}}) = (0.800, 0.500)$, (iii) $(P_{\mathrm{ILI}}, P_{\mathrm{fail}}) = (0.004, 0.000)$, and (iv) $(P_{\mathrm{ILI}}, P_{\mathrm{fail}}) = (0.800, 0.000)$. **B:** Daily number of new infections and daily number of new positive PCR tests in Scenario C. The quadrant settings are identical to **A**.

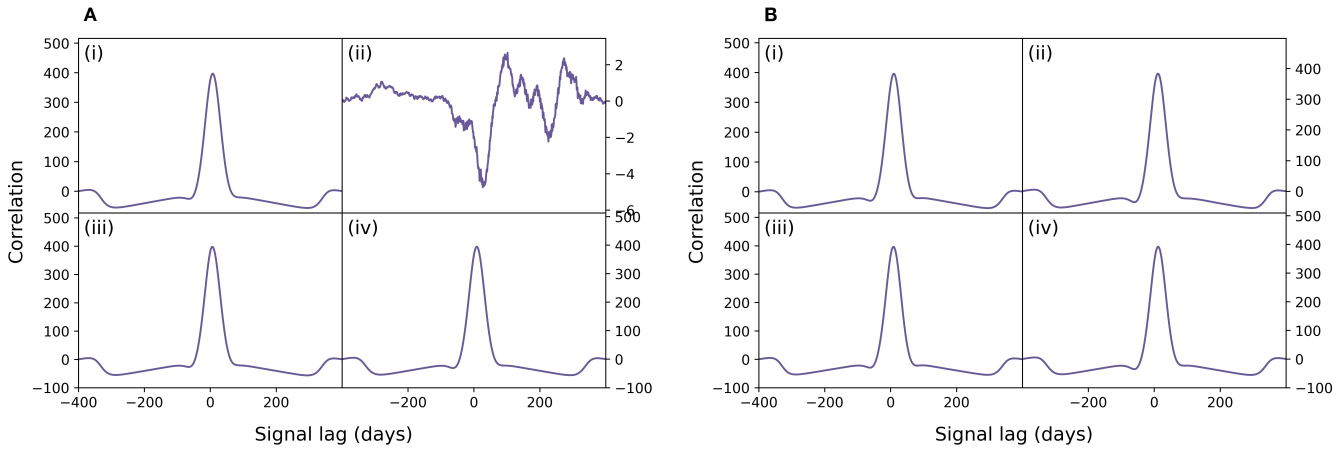


**Fig. S8. A:** Cross correlation function evaluated over the daily number of new infections and the daily number of new positive RATs in Scenario C. The cross correlation function in quadrant (ii) cannot be resolved due to an essentially constant signal of daily positive RATs in Figure S7**A**(ii). The quadrant settings are (i) $(P_{\mathrm{ILI}}, P_{\mathrm{fail}}) = (0.004, 0.500)$, (ii) $(P_{\mathrm{ILI}}, P_{\mathrm{fail}}) = (0.800, 0.500)$, (iii) $(P_{\mathrm{ILI}}, P_{\mathrm{fail}}) = (0.004, 0.000)$, and (iv) $(P_{\mathrm{ILI}}, P_{\mathrm{fail}}) = (0.800, 0.000)$. **B:** Cross correlation function evaluated over the daily number of new infections and the daily number of new positive PCR tests in Scenario C$^{*}$. The quadrant settings are identical to **A**.

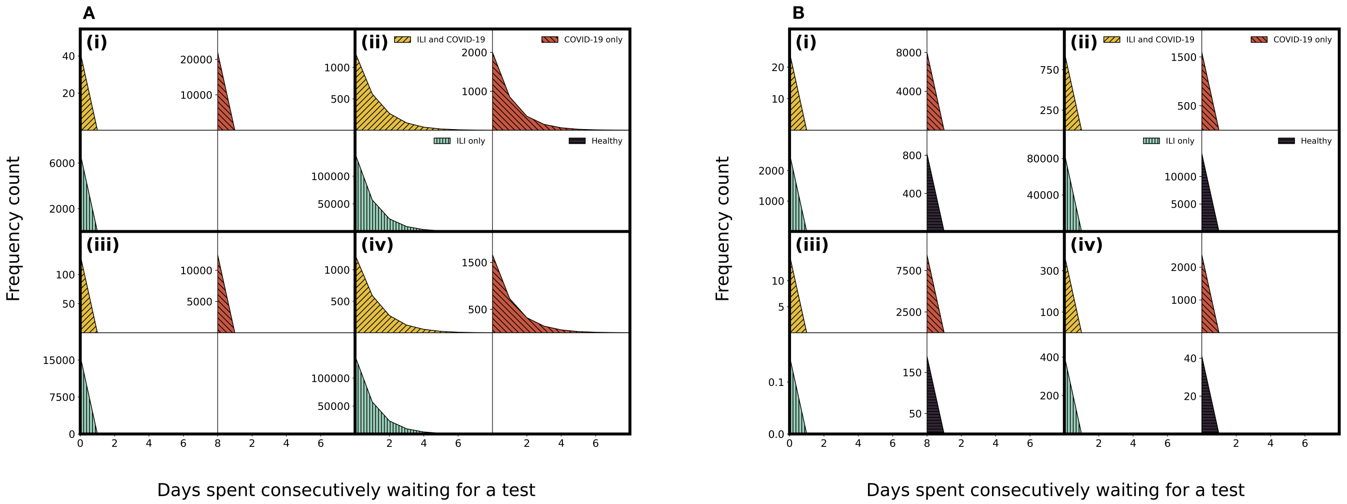


**Fig. S9. A:** Waiting time distribution of RATs for agents of any infection status in Scenario C. Each quadrant corresponds to a setting of $(P_{\mathrm{ILI}}, P_{\mathrm{fail}})$ and is itself divided into sub-quadrants to display each infection status. A sub-quadrant is blank if no RAT was ever taken by an agent with the corresponding infection status. The quadrant settings are (i) $(P_{\mathrm{ILI}}, P_{\mathrm{fail}}) = (0.004, 0.500)$, (ii) $(P_{\mathrm{ILI}}, P_{\mathrm{fail}}) = (0.800, 0.500)$, (iii) $(P_{\mathrm{ILI}}, P_{\mathrm{fail}}) = (0.004, 0.000)$, and (iv) $(P_{\mathrm{ILI}}, P_{\mathrm{fail}}) = (0.800, 0.000)$. **B:** Waiting time distribution of PCR tests for agents of any infection status in Scenario C. Each quadrant corresponds to a setting of $(P_{\mathrm{ILI}}, P_{\mathrm{fail}})$ and is itself divided into sub-quadrants to display each infection status. The quadrant settings are identical to **A**.

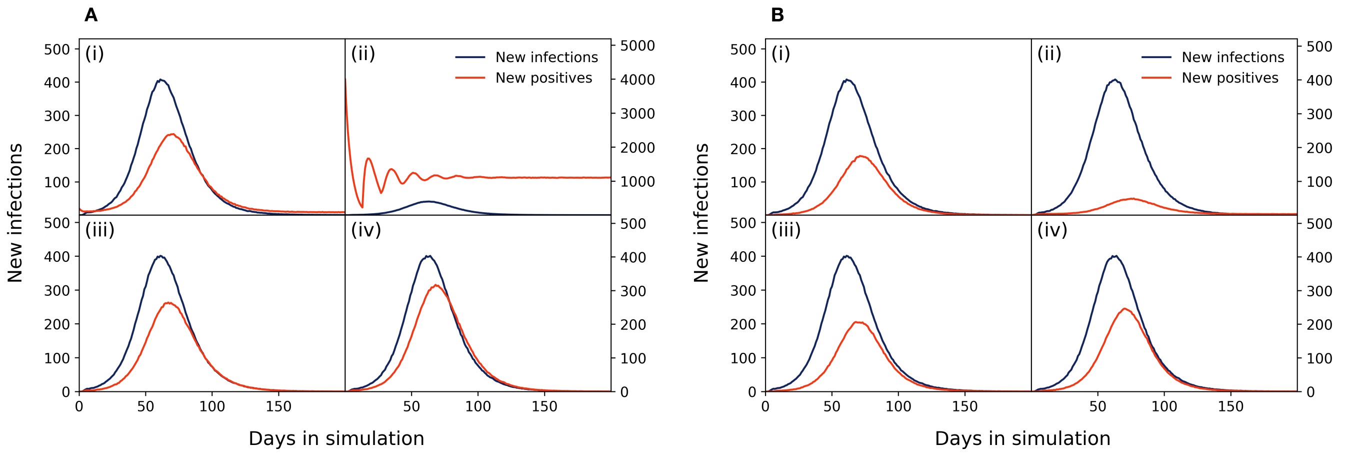


**Fig. S10.** **A:** Daily number of new infections and daily number of new positive RATs in Scenario C$^*$. Oscillations arise in (ii) due to initial synchronization of positive re-test delays. The quadrant settings are (i) $(P_{\mathrm{ILI}}, P_{\mathrm{fail}}) = (0.004, 0.500)$, (ii) $(P_{\mathrm{ILI}}, P_{\mathrm{fail}}) = (0.800, 0.500)$, (iii) $(P_{\mathrm{ILI}}, P_{\mathrm{fail}}) = (0.004, 0.000)$, and (iv) $(P_{\mathrm{ILI}}, P_{\mathrm{fail}}) = (0.800, 0.000)$. **B:** Daily number of new infections and daily number of new positive PCR tests in Scenario C$^*$. The quadrant settings are identical to **A**.

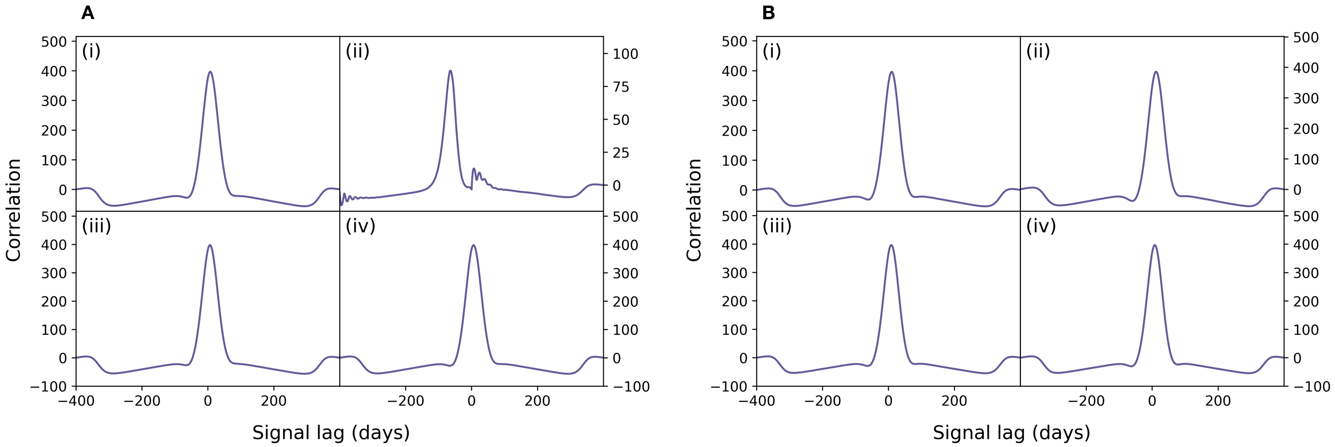


**Fig. S11. A:** Cross correlation function evaluated over the daily number of new infections and the daily number of new positive RATs in Scenario C$^*$. The cross correlation function in quadrant (ii) cannot be resolved due to the presence of strong oscillations in the signal of daily positive RATs in Figure S10**A**(ii). The quadrant settings are (i) $(P_{\mathrm{ILI}}, P_{\mathrm{fail}}) = (0.004, 0.500)$, (ii) $(P_{\mathrm{ILI}}, P_{\mathrm{fail}}) = (0.800, 0.500)$, (iii) $(P_{\mathrm{ILI}}, P_{\mathrm{fail}}) = (0.004, 0.000)$, and (iv) $(P_{\mathrm{ILI}}, P_{\mathrm{fail}}) = (0.800, 0.000)$. **B:** Cross correlation function evaluated over the daily number of new infections and the daily number of new positive PCR tests in Scenario C$^*$. The quadrant settings are identical to **A**.

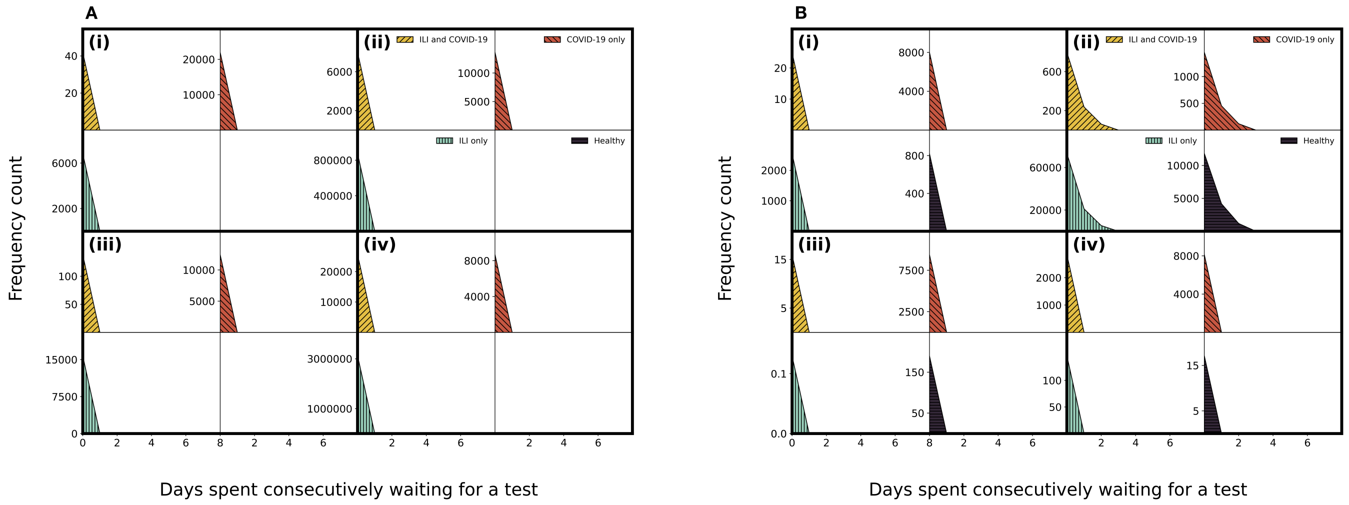


**Fig. S12. A:** Waiting time distribution of RATs for agents of any infection status in Scenario C$^*$. Each quadrant corresponds to a setting of $(P_{\mathrm{ILI}}, P_{\mathrm{fail}})$ and is itself divided into sub-quadrants to display each infection status. A sub-quadrant is blank if no RAT was ever taken by an agent with the corresponding infection status. The quadrant settings are (i) $(P_{\mathrm{ILI}}, P_{\mathrm{fail}}) = (0.004, 0.500)$, (ii) $(P_{\mathrm{ILI}}, P_{\mathrm{fail}}) = (0.800, 0.500)$, (iii) $(P_{\mathrm{ILI}}, P_{\mathrm{fail}}) = (0.004, 0.000)$, and (iv) $(P_{\mathrm{ILI}}, P_{\mathrm{fail}}) = (0.800, 0.000)$. **B:**. Waiting time distribution of PCR tests for agents of any infection status in Scenario C$^*$. Each quadrant corresponds to a setting of $(P_{\mathrm{ILI}}, P_{\mathrm{fail}})$ and is itself divided into sub-quadrants to display each infection status. The quadrant settings are identical to **A**.
